# Assessing the Feasibility of a Web-based Outcome Measurement System in Child and Adolescent Mental Health Services: myHealthE (MHE) a Randomised Controlled Feasibility Pilot Study

**DOI:** 10.1101/2021.10.07.21264418

**Authors:** Anna C Morris, Zina Ibrahim, Margaret Heslin, Omer S Moghraby, Argyris Stringaris, Ian M Grant, Lukasz Zalewski, Megan Pritchard, Robert Stewart, Matthew Hotopf, Andrew Pickles, Richard J B Dobson, Emily Simonoff, Johnny Downs

## Abstract

**Background:** Interest in internet-based patient reported outcome measure (PROM) collection is increasing. The NHS myHealthE (MHE) web-based monitoring system was developed to address the limitations of paper-based PROM completion. MHE provides a simple and secure way for families accessing Child and Adolescent Mental Health Services to report clinical information and track their child’s progress. This study aimed to assess whether MHE improves the completion of the Strengths and Difficulties Questionnaire (SDQ) compared with paper collection. Secondary objectives were to explore caregiver satisfaction and application acceptability.

**Methods:** A twelve-week single-blinded randomised controlled feasibility pilot trial of MHE was conducted with 196 families accessing neurodevelopmental services in south London to examine whether electronic questionnaires are completed more readily than paper-based questionnaires over a 3-month period. Follow up process evaluation phone calls with a subset (n=8) of caregivers explored system satisfaction and usability.

**Results:** MHE group assignment was significantly associated with an increased probability of completing an SDQ-P in the study period (adjusted hazard ratio, (HR) 12.1, 95% CI 4.7-31.0; *p*= <0.001). Of those caregivers’ who received the MHE invitation (n=68) 69.1% completed an SDQ using the platform compared to 8.8% in the control group (n=68). The system was well received by caregivers, who cited numerous benefits of using MHE, for example, real time feedback and ease of completion.

**Conclusions:** MHE holds promise for improving PROM completion rates. Research is needed to refine MHE, evaluate large scale MHE implementation, cost effectiveness and explore factors associated with differences in electronic questionnaire uptake.

**Key practitioner messages:**

- Patient reported outcome measures (PROMs) are considered an important tool for measuring treatment success and outcomes in healthcare systems.
- Adherence to routine PROM guidance in Child and Adolescent Mental Health Services (CAMHS) remains low, largely driven by limitations associated with paper-based data collection.
- Paperless monitoring systems (i.e., digital) as an alternative to traditional outcome measure delivery and collection are growing in healthcare settings.
- Remote questionnaire completion using the myHealthE (MHE) system is feasible and acceptable to caregivers of children accessing CAMHS in South London, yielding a 60.3 % increase in Strengths and Difficulties questionnaire reporting compared to standard practice.
- More research is required to understand whether MHE implementation affords similar improvements in remote PROM completion at scale and whether electronic questionnaire uptake is equal for different socio-demographic and clinical populations.

## Introduction

Patient-reported outcome measures (PROMs) enable standardised and direct collection of a patient’s perceived health status (Devlin & Appleby, 2010; NHS England, 2019). Used routinely, PROMs are recognised as a clinically valuable method to measure patient or caregiver rated symptoms, assess intervention success, and encourage shared patient and practitioner communication and decision making (Carlier, Meuldijk, Van Vliet et al., 2012; Lambert, Whipple, Hawkins et al., 2003; Soreide & Soreide, 2013). Child and Adolescent Mental Health Services (CAMHS) in England are encouraged to collect information about young people’s presenting problems at entry to CAMHS and again within six months of receiving treatment (Department of Health (DoH), 2004 & DoH, 2015) using patient PROMs. However, audit and survey studies demonstrate low guideline adherence, suggesting that CAMHS struggle to implement PROMs (Batty, Moldavsky, Foroushani et al., 2013; Hall, Moldavsky, Baldwin et al. 2013; Johnston & Gowers 2005). Recent research investigating the electronic health records of 28,000 patients accessing CAMH services across South London identified paired use of the Strengths and Difficulties Questionnaire (SDQ; Goodman, 1997), a readily available behavioural screening tool at baseline and follow up in only 8% of these (Morris, Macdonald, Moghraby et al., 2020) and as few as 1% within specific clinical groups, namely attention deficit hyperactivity disorder (ADHD) (Cruz, Simonoff, McGough et al., 2015).

Data collection using traditional paper questionnaires is associated with several time- and resource-intensive steps, including printing, postage and processing returned outcome measures. Although paper questionnaires are practical and easy to complete, already-burdened clinicians struggle with the administrative effort required to capture paper-based questionnaires (Boswell, Kraus, Miller et al., 2015; Johnston and Gowers, 2005; Hall, Taylor, Moldavsky et al, 2014). Response data are also easily comprised, for example, users can omit questions, select multiple responses per item, and mark outside the questions tick box margins, leading to missing or unusable data (Ebert, Huibers, Christensen et al., 2018).

A rapid rise in internet use has paved the way for electronic questionnaires (Lyon, Lewis, Boyd et al., 2016). Electronic PROMs (ePROMs) are reported to be less time consuming (Cella, Hanh, Jensen et al., 2015), require fewer administrative duties (Black, 2013; Coons, Eremenco, Lundy, et al., 2015; Eremenco, Coons, Paty et al., 2014), cost less (Zuidgeest, Hendriks, Koopman et al., 2011) and evoke more honest (Black & Ponirakis, 2000) and less erroneous responses by prompting patients to respond to all items within a questionnaire and only provide one response per question (Dillon, Pirie, Rice et al., 2014; Jamison, Raymond, Levine et al., 2001; Coons et al, 2015; Eremenco et al, 2014).

Feasibility trials of web-based monitoring systems report positive outcomes relating to patient engagement, satisfaction, and clinical value (Schepers, Sint Nicolaas, Maurice-Stam et al., 2017; Barthel, Fischer, Nolte et al., 2016; Ashley, Jones, Thomas et al., 2013, Nordan, Blanchfield, Niazi et al., 2018). However, less research is available on the development and application of ePROM systems in CAMHS. Interviews with mental health service users demonstrate positive attitudes toward the use of technology to assist traditional care (Borzekowski, Leith, Medoff et al., 2009). However, patients have highlighted barriers to web-based portal acceptability, including computer literacy, perceived usefulness, suitability, confidentiality, feedback and the effect application use has on their capacity to manage their condition and therapeutic relationships (Niazkhani, Toni, Cheshmekaboodi et al., 2020).

The myHealthE (MHE) system was built to enable remote PROM monitoring in CAMHS. This system aims to automate the communication, delivery and collection of ePROMs at predefined post-treatment periods, providing caregivers with a safe and engaging way to share clinically relevant information about their child with their allocated care team with minimal human input. MHE architecture, development and implementation methodology have been described previously (Morris, Ibrahim, Moghraby, 2021). Novel healthcare applications require feasibility and acceptability testing to ensure that the technology is understandable and can be used successfully by the target end-user in real-world clinical surroundings before conducting a large-scale system evaluation (Steele Gray, Gill, Khan et al., 2016a). As described in our protocol [(ISRCTN) 22581393], the primary purpose of the trial was to understand whether MHE use should be assessed in CAMHS on a wider scale. Therefore, we conducted a feasibility pilot study of MHE to evaluate whether introducing MHE enhanced completion of PROM questionnaires over the course of CAMHS treatment compared to standard data collection procedures, as measured by a count of how many electronic questionnaires were completed relative to paper questionnaires over a three-month period. Secondly, we aimed to assess caregiver satisfaction with the system via individual caregiver phone consultations. Given resource constraints we were unable to assess the economic benefit of MHE compared to standard data acquisition as per our protocol. We hypothesised that MHE implementation would afford a substantial increase in completed standardised caregiver-reported follow up data and caregiver satisfaction with CAMHS services compared to routine data collection.

## Methods

### Design

The current study comprised a single-blinded parallel group feasibility pilot randomised control trial (RCT) of MHE. Outcome, sociodemographic and service level data were obtained from the Clinical Record Interactive Search (CRIS) system. CRIS contains de-identified medical record history from the South London and Maudsley (SLaM) National Health Service Foundation Trust, one of Europe’s largest mental health care organisations providing services to over 34,400 children and adolescents between the 1^st^ of January 2008 and 1^st^ December 2018 (Stewart, Soremekun, Perera et al., 2009; Perera, Broadbent, Callard et al., 2016; Downs, Ford, Stewart et al., 2019). This research tool was established by SLaM’s National Institute of Health Research Biomedical Research Centre (NIHR BRC) in 2008, to enable information retrieval for the purpose of approved research (Fernandes, Cloete, Broadbent et al, 2013). Comprehensive electronic health record (EHR) information is available for SLaM services from 2006.

### Setting and participants

The trial was conducted at Kaleidoscope, a community paediatric and children’s and young people’s mental health centre based in Lewisham, South London between the 11th of February 2019 and the 14th of May 2019. Eligible participants were caregivers of active CAMHS patients aged between 4-18 years old with a diagnosis of autism spectrum disorder (ASD). Patients were under the care of Lewisham Neuro-developmental Team and had at least one SDQ present in their EHR. Caregivers were recruited if they had available contact details (mobile phone number and/or email address) in their child’s EHR. The MHE data collection process was directly comparable to current paper-based practice, except for its electronic basis as such, the system only collects data which is ordinarily requested from families by their treating clinical team and its use does not instigate change in patient care. Therefore, caregivers did not have to provide informed consent to participate in this trial. Instead, caregivers could choose to opt-out via email or phone call to the trial research assistant (ACM). Recruitment was achieved through SLaM EHR screening. A SQL script developed and implemented by a senior member of the SLaM Clinical Systems Team automatically generated an extract of eligible patients which was then provided to the research team. Subsequently, computerised condition allocation and simple randomisation was performed to assign eligible caregivers to either receive PROM outcome monitoring as usual (MAU; control group) or enrolment to the MHE platform (intervention group) on a 1:1 basis. Clinicians were blinded to condition allocation, whereby they were not informed which patients on their case load had been allocated to received MHE or MAU.

### Measures, sociodemographic and clinical characteristics

The primary outcome variable was the number follow-up caregiver reported SDQ (SDQ-P; electronic vs paper SDQ-P) forms completed over the three-month observation period. The SDQ-P (Appendix 1) is a structured 25-item questionnaire screening for symptoms of childhood emotional and behavioural psychopathology (Goodman, 1997). SLaM holds a sub-licence to use the SDQ to support clinical service via NHS Digital Copyright Licensing Service. It is current clinical practice to collect SDQ-P for young people, either by post before their first face-to-face meeting, or on site during a clinical appointment to inform their baseline assessment and again six months after starting treatment or upon discharge from CAMHS. Other variables extracted from CRIS are presented in online supplement 1.

### Process evaluation: usability testing

To evaluate usability, from a subset of caregivers randomly assigned to MHE a convenience sample of 6 caregivers who had engaged with MHE and 2 caregivers who had not were contacted by telephone. These caregivers were asked to complete the System Usability Scale (SUS; Brooke, 1996) to examine subjective usability. SUS comprises 10 statements reported on a 5-point Likert scale ranging from strongly disagree to strongly agree. The total score is presented as a figure from 0 to 100, with a greater score reflecting higher usability. Mean SUS score was computed and ranked using Bangor, Kortum and Miller’s (2008) acceptability scale defined as ‘not acceptable’, ‘marginal’ and ‘acceptable’. Following administration of the SUS, caregivers were invited to ask questions about the platform or provide any further comments about their experience of using MHE.

### Sample size

The current trial aimed to inform the development of a larger, adequately powered RCT by providing precise estimates of acceptability and feasibility, in addition to outcome variability. A threshold of clinical significance was decided *a priori* to be 15% between MAU and MHE groups for SDQ-P completion within three-months, based on consensus from Kaleidoscope staff and previous research indicating an expected baseline completion rate of 8% SDQ-P in the control group (Morris et al, 2020). For a fixed sample size design, the sample size required to achieve a power of 1-β = 0.80 for the two-tailed chi-square test at level α = 0.05, under the prior assumptions, was 2 × 91 = 182 on a 1:1 allocation ratio. The power calculation was carried out using Gpower 3.1.7. To increase power and reduce the risk of chance imbalance between MHE and non-MHE groups, we followed recent guidance on covariate adjustment within RCTs of moderate sample size [https://trialsjournal.biomedcentral.com/articles/10.1186/1745-6215-15-139], and included in our survival analyses, several factors which could have potential influence on PROM completion (Morris et al, 2020).

### Intervention and Procedure

Figure 1 provides an overview and description of the MHE data flow. All caregivers of active Lewisham Neurodevelopmental Team patients were contacted by letter to inform them of potential changes to the way Lewisham CAMHS gather clinical information about their patients (i.e. electronic rather than paper questionnaires) and provided with an patient information sheet and MHE information leaflet (Appendix 7a and 7b). After group assignment, caregivers allocated to receive MHE were contacted with an invitation to set up a personalised web-portal and complete an SDQ-P (caregivers were enrolled to the trial irrespective of whether they registered their MHE account if baseline SDQ information was present in their child’s EHRs). Caregivers who did not register were prompted every week to enrol and complete an SDQ-P. Once an online questionnaire was completed, they were contacted monthly to provide new SDQ data. In the control group caregivers were requested to complete paper SDQ-P face-to-face or by post according to clinician discretion. Apart from electronic SDQ-P completion for the intervention group, treatment remained the same for all participants. Information collected through MHE was stored in the child’s EHR and managed in the same way as all other confidential information. SDQ-P data was checked daily by ACM and promptly entered to the patient’s EHR. Post intervention, all participants received a letter thanking them for their participation.

**Figure 1.**
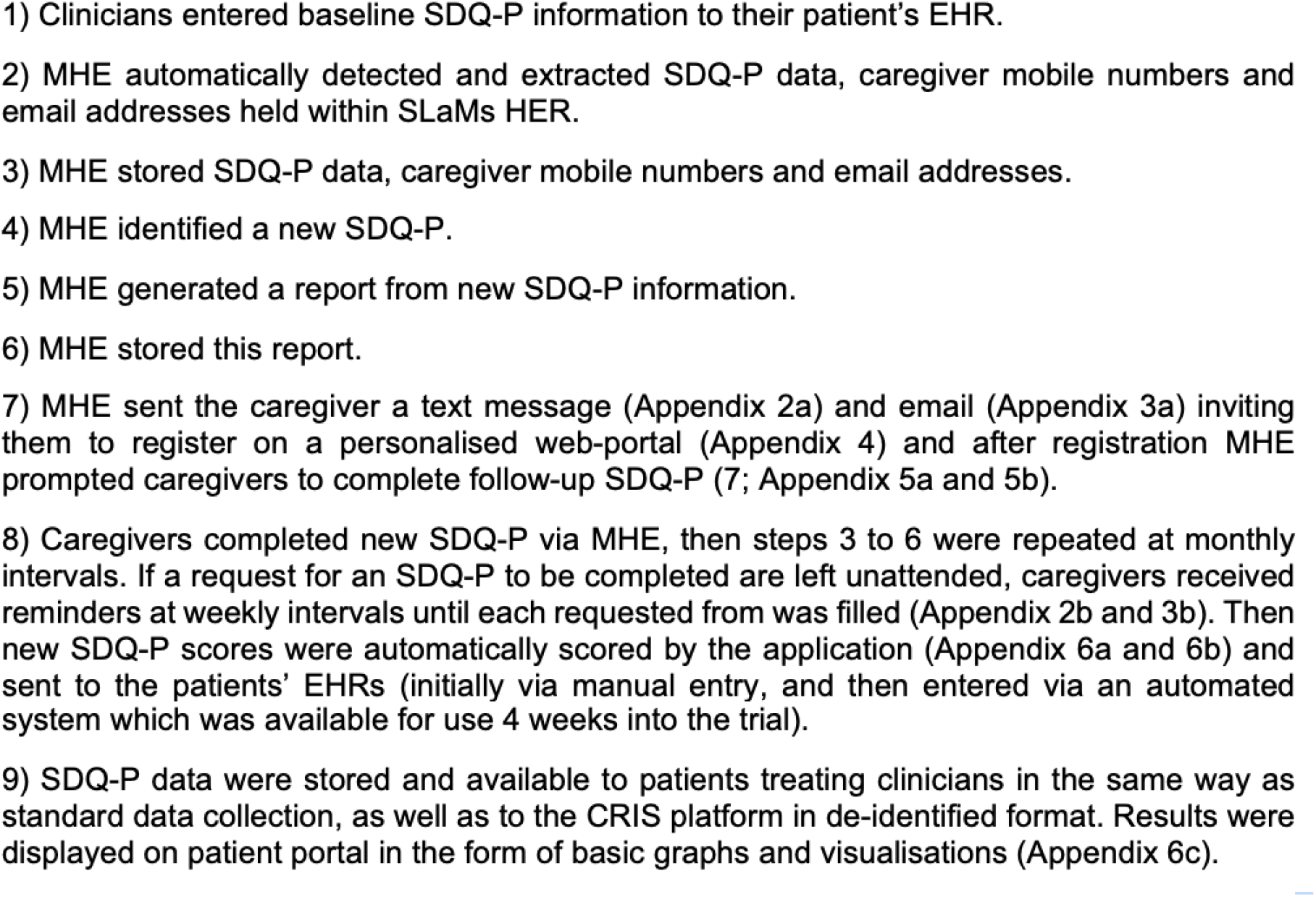
myHealthE data flow-diagram - contact the corresponding author to request access to this image.

### Strategy for analysis

All analyses were conducted using STATA version 14 (StataCorp, 2015). Analyses were conducted to determine differences in the primary outcome, namely paper relative to electronic follow-up SDQ-P completion. Analysis was performed subject to intention-to-treat like principles (intention-to-contact), whereby all participants were analysed according to their initially assigned intervention arm, irrespective of protocol adherence or deviations. Cox regression was used to examine the relationship between monitoring as usual vs MHE group assignment and SDQ-P completion rates. Using a Kaplan-Meier curve, we checked whether group assignment (as predictor) satisfied the proportional hazards assumption. Our first analysis examined the association between treatment group only and SDQ-P completion. The second model adjusted for demographic and clinical covariates captured in this trial. An inverse Kaplan-Meier curve was plotted to visualise the probability of SDQ-P completion, comparing caregivers who completed electronic and paper SDQ-P. For the intervention group the MHE website–SDQ-P completion conversion rate was reported as a percentage by measuring the number of caregivers that register on MHE and subsequently completed a follow-up SDQ-P.

## Results

### Enrolment and baseline characteristics

Within study participant flow and data collection rates are provided in Figure 2. A total of 342 caregivers were screened for eligibility of which (n=196) met the inclusion criteria. During eligibility screening caregiver contact information was often missing or located in an area of the patients’ EHRs different from expected, therefore manual contact detail collection was carried out to enable digital communication via MHE. Caregivers were enrolled and randomly assigned to the intervention group (MHE n=98) and the control group (MAU n=98). Of caregivers assigned to MHE and MAU 30 (36.3%) did not receive notifications from MHE. The conversion rate from account registration to SDQ completion was 98% (47/48). Online supplement 2 outlines account registration issues and opt-out preferences reported by caregivers.

**Figure 2.**
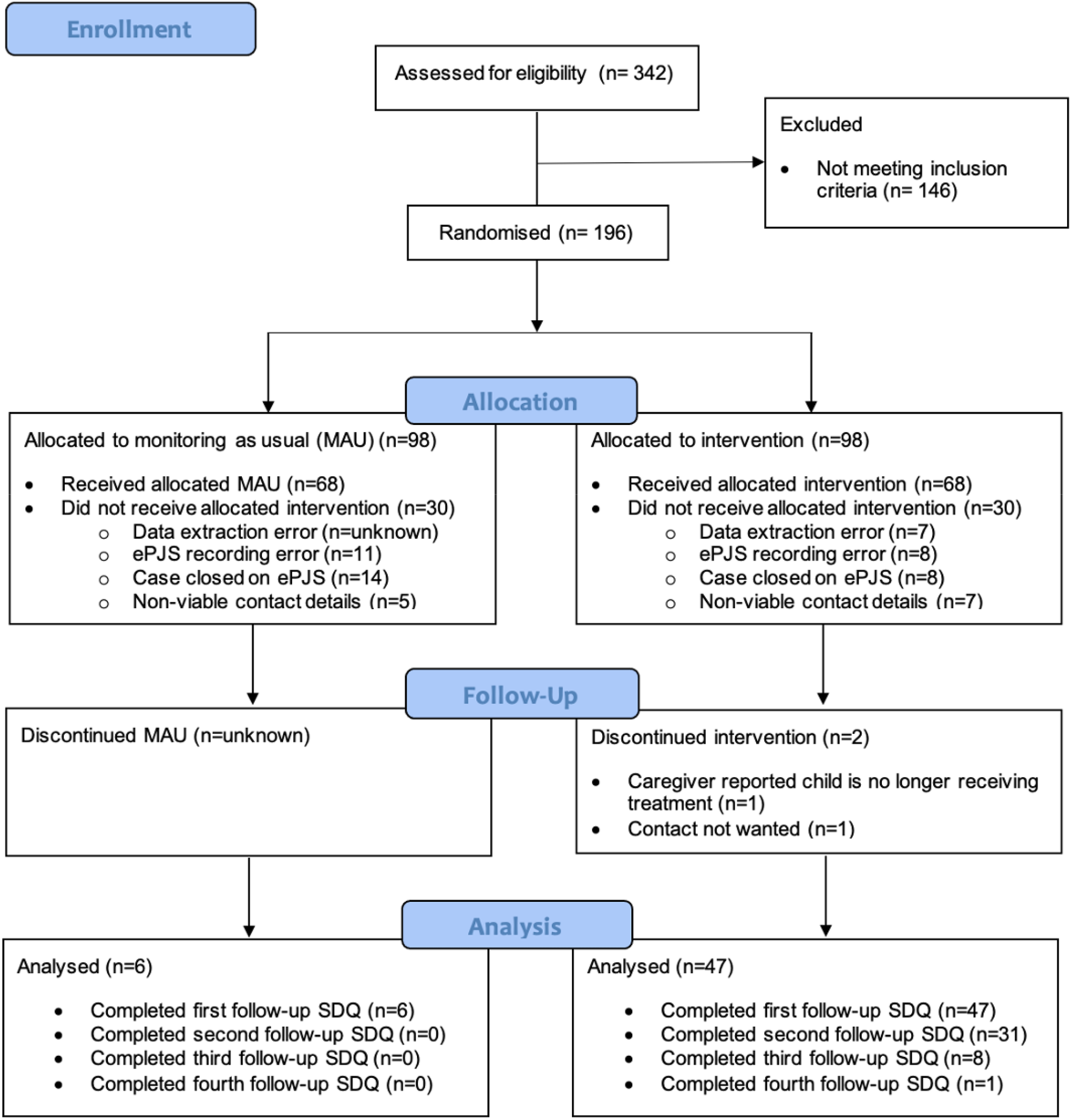
Consort diagram presenting recruitment and rate of data collection for MHE and MAU.

Table 1. presents sociodemographic and service characteristics for the whole sample. Participants were ethnically diverse, predominantly male and at the older end of the age range accepted by CAMHS.

**Table 1.**
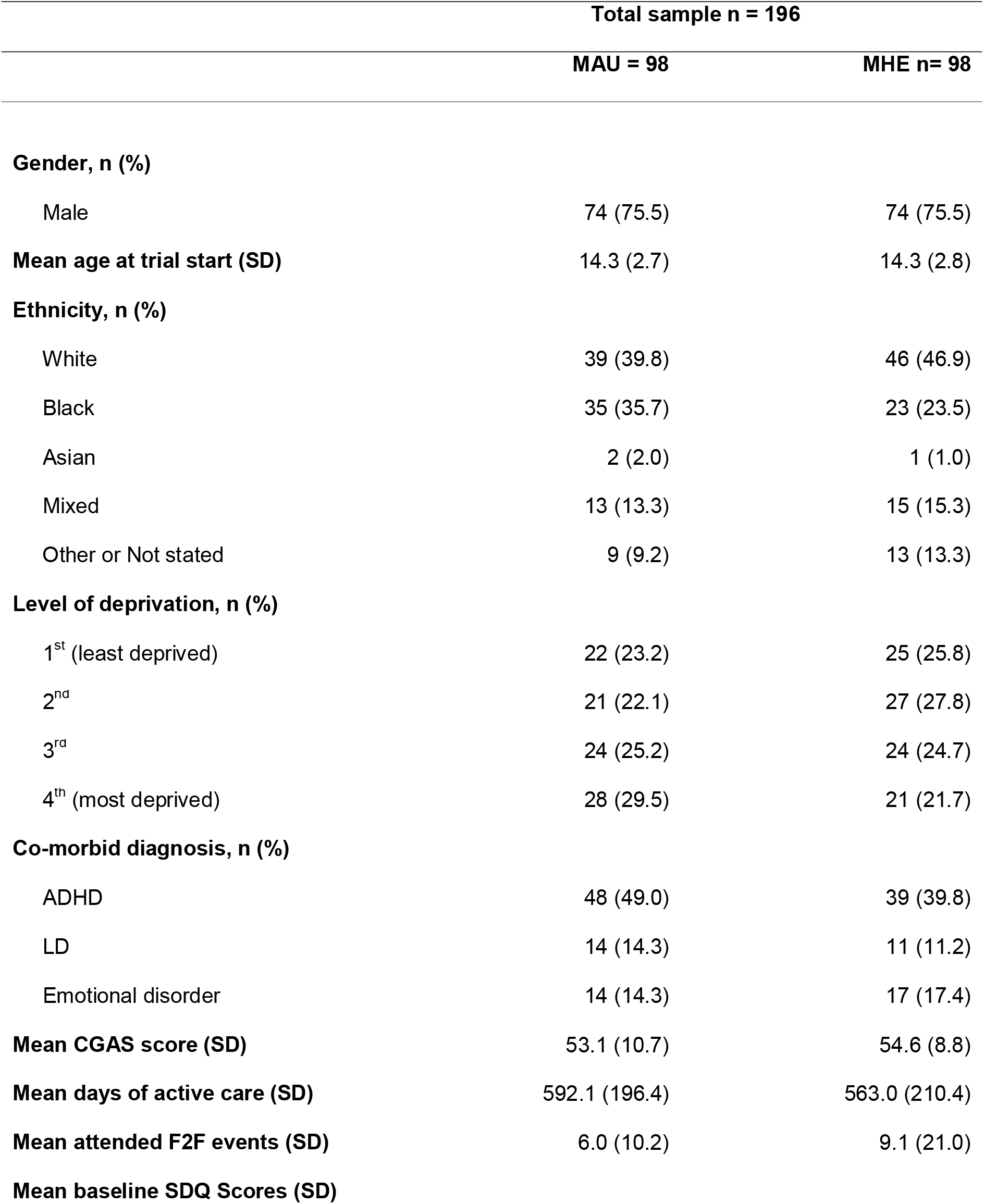
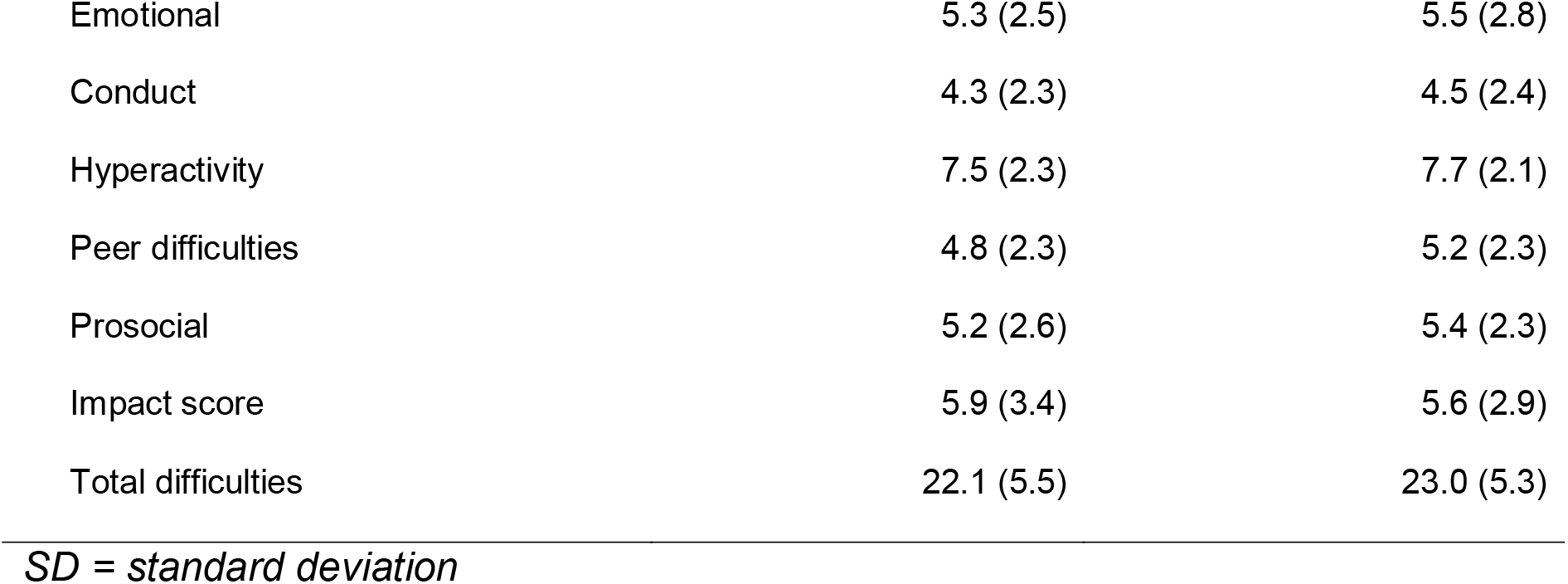
Baseline patient and clinical characteristics of MHE vs MAU following randomisation (n=196).

|  | Total sample n = 196 |  |
| --- | --- | --- |
|  | MAU = 98 | MHE n= 98 |
| **Gender, n (%)** |  |  |
| Male | 74 (75.5) | 74 (75.5) |
| **Mean age at trial start (SD)** | 14.3 (2.7) | 14.3 (2.8) |
| **Ethnicity, n (%)** |  |  |
| White | 39 (39.8) | 46 (46.9) |
| Black | 35 (35.7) | 23 (23.5) |
| Asian | 2 (2.0) | 1 (1.0) |
| Mixed | 13 (13.3) | 15 (15.3) |
| Other or Not stated | 9 (9.2) | 13 (13.3) |
| **Level of deprivation, n (%)** |  |  |
| 1<sup>st</sup> (least deprived) | 22 (23.2) | 25 (25.8) |
| 2<sup>nd</sup> | 21 (22.1) | 27 (27.8) |
| 3<sup>rd</sup> | 24 (25.2) | 24 (24.7) |
| 4<sup>th</sup> (most deprived) | 28 (29.5) | 21 (21.7) |
| **Co-morbid diagnosis, n (%)** |  |  |
| ADHD | 48 (49.0) | 39 (39.8) |
| LD | 14 (14.3) | 11 (11.2) |
| Emotional disorder | 14 (14.3) | 17 (17.4) |
| **Mean CGAS score (SD)** | 53.1 (10.7) | 54.6 (8.8) |
| **Mean days of active care (SD)** | 592.1 (196.4) | 563.0 (210.4) |
| **Mean attended F2F events (SD)** | 6.0 (10.2) | 9.1 (21.0) |
| **Mean baseline SDQ Scores (SD)** |  |  |
| Emotional | 5.3 (2.5) | 5.5 (2.8) |
| Conduct | 4.3 (2.3) | 4.5 (2.4) |
| Hyperactivity | 7.5 (2.3) | 7.7 (2.1) |
| Peer difficulties | 4.8 (2.3) | 5.2 (2.3) |
| Prosocial | 5.2 (2.6) | 5.4 (2.3) |
| Impact score | 5.9 (3.4) | 5.6 (2.9) |
| Total difficulties | 22.1 (5.5) | 23.0 (5.3) |

*SD = standard deviation*

### Electronic vs paper SDQ-P collection

During the trial 47 caregivers [47.9% of intention-to-contact (total n=98), 69.1% of actually contacted (total n=68)] registered an account on the MHE platform and completed at least one follow-up SDQ-P. In the corresponding timeframe 6 (intention to contact = 6% (n=98) and actually contacted = 8.8% (n=68) caregivers assigned to receive MAU completed at least one follow-up SDQ-P. Second follow-up was due for 43 of the MHE cohort by the end of the study period (at least one month had elapsed since completing their first online SDQ-P) and of these 31 caregivers completed this (72%). Overall, 87 follow-up SDQ-Ps were completed via the MHE platform: Figure 3. provides a breakdown of SDQ-P completion within each 7-day notification reminder period.

**Figure 3.**
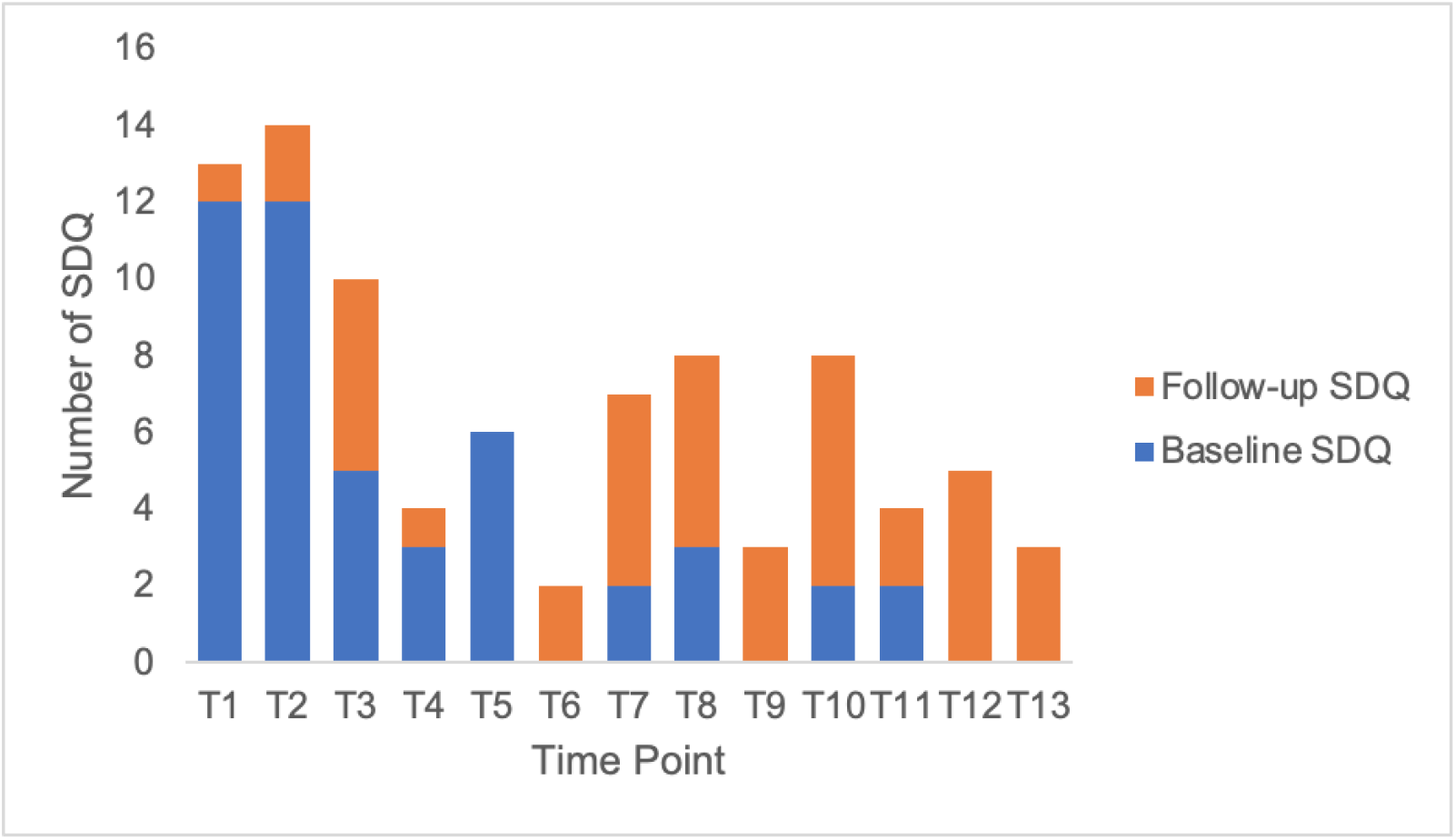
Baseline and follow-up SDQ completion within each 7-day notification period.

The ITC Cox regression models are presented in Table 2, and graphically depicted in Figure 4. MHE group assignment was significantly associated with an increased probability of completing an SDQ-P in the study period (adjusted hazard ratio, (HR) 12.1, 95% CI 4.7-31.0; *p*= <0.001). This was observed after controlling for potentially confounding socio-demographic characteristics and clinical factors including, gender, age at the start if the trial, baseline CGAS and SDQ profiles, co-morbid ADHD, learning disability, and emotional disorders as well as number of days of active care and attended face-to-face events. No significant interaction was found between ethnic status (white and non-white ethnic groups) and SDQ-P completion by group.

**Table 2.**
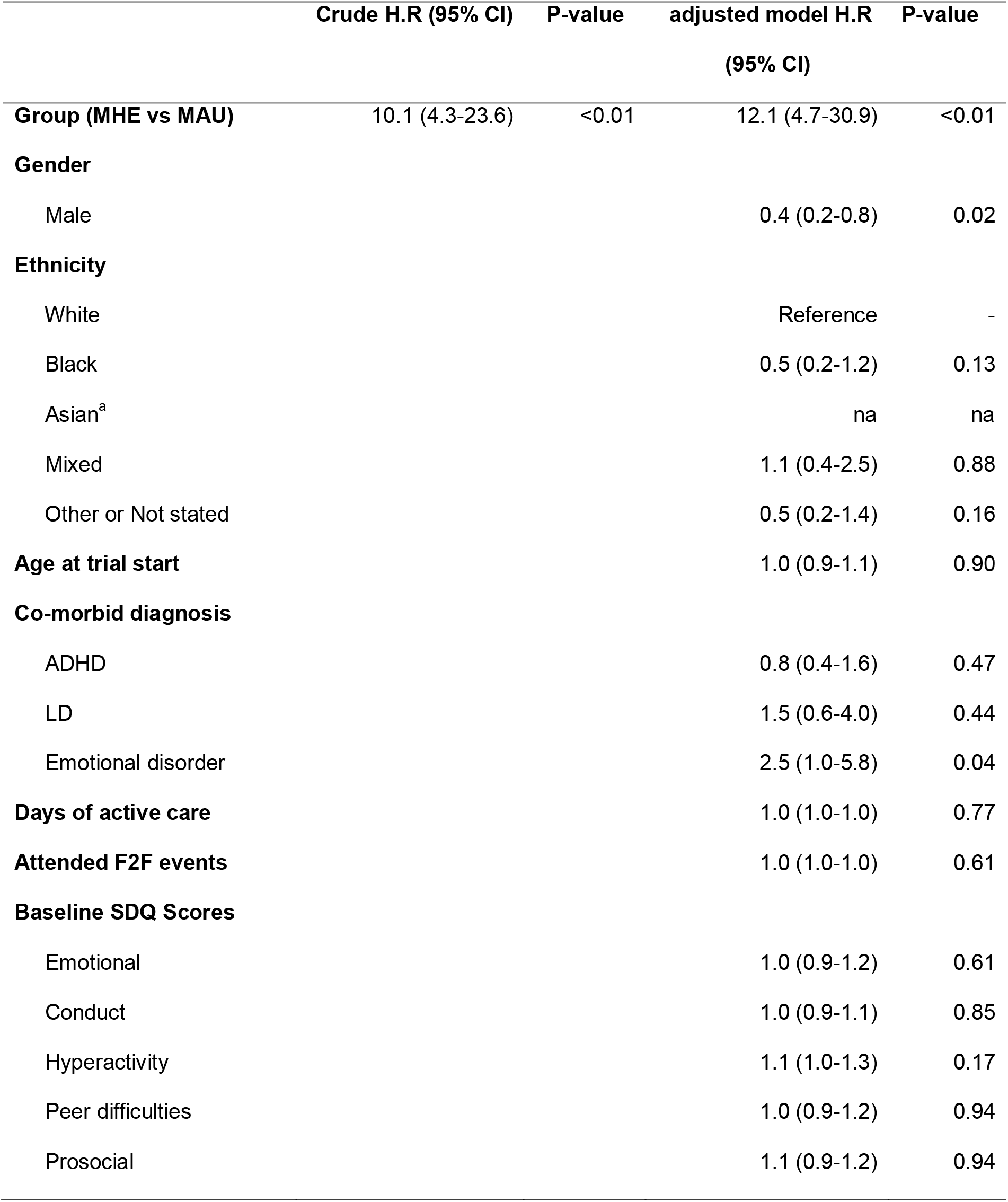
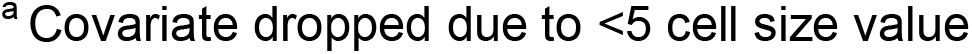
An Intention to contact Cox-regression analysis of the relationship between electronic compared to paper-based SDQ-P assignment and SDQ-P completion rates (n=195), adjusted model taking into account participant characteristics.

|  | Crude H.R (95% CI) | P-value | adjusted model H.R (95% CI) | P-value |
| --- | --- | --- | --- | --- |
| **Group (MHE vs MAU)** | 10.1 (4.3-23.6) | <0.01 | 12.1 (4.7-30.9) | <0.01 |
| **Gender** |  |  |  |  |
| Male |  |  | 0.4 (0.2-0.8) | 0.02 |
| **Ethnicity** |  |  |  |  |
| White |  |  | Reference | - |
| Black |  |  | 0.5 (0.2-1.2) | 0.13 |
| Asian<sup>a</sup> |  |  | na | na |
| Mixed |  |  | 1.1 (0.4-2.5) | 0.88 |
| Other or Not stated |  |  | 0.5 (0.2-1.4) | 0.16 |
| **Age at trial start** |  |  | 1.0 (0.9-1.1) | 0.90 |
| **Co-morbid diagnosis** |  |  |  |  |
| ADHD |  |  | 0.8 (0.4-1.6) | 0.47 |
| LD |  |  | 1.5 (0.6-4.0) | 0.44 |
| Emotional disorder |  |  | 2.5 (1.0-5.8) | 0.04 |
| **Days of active care** |  |  | 1.0 (1.0-1.0) | 0.77 |
| **Attended F2F events** |  |  | 1.0 (1.0-1.0) | 0.61 |
| **Baseline SDQ Scores** |  |  |  |  |
| Emotional |  |  | 1.0 (0.9-1.2) | 0.61 |
| Conduct |  |  | 1.0 (0.9-1.1) | 0.85 |
| Hyperactivity |  |  | 1.1 (1.0-1.3) | 0.17 |
| Peer difficulties |  |  | 1.0 (0.9-1.2) | 0.94 |
| Prosocial |  |  | 1.1 (0.9-1.2) | 0.94 |

**Figure 4.**
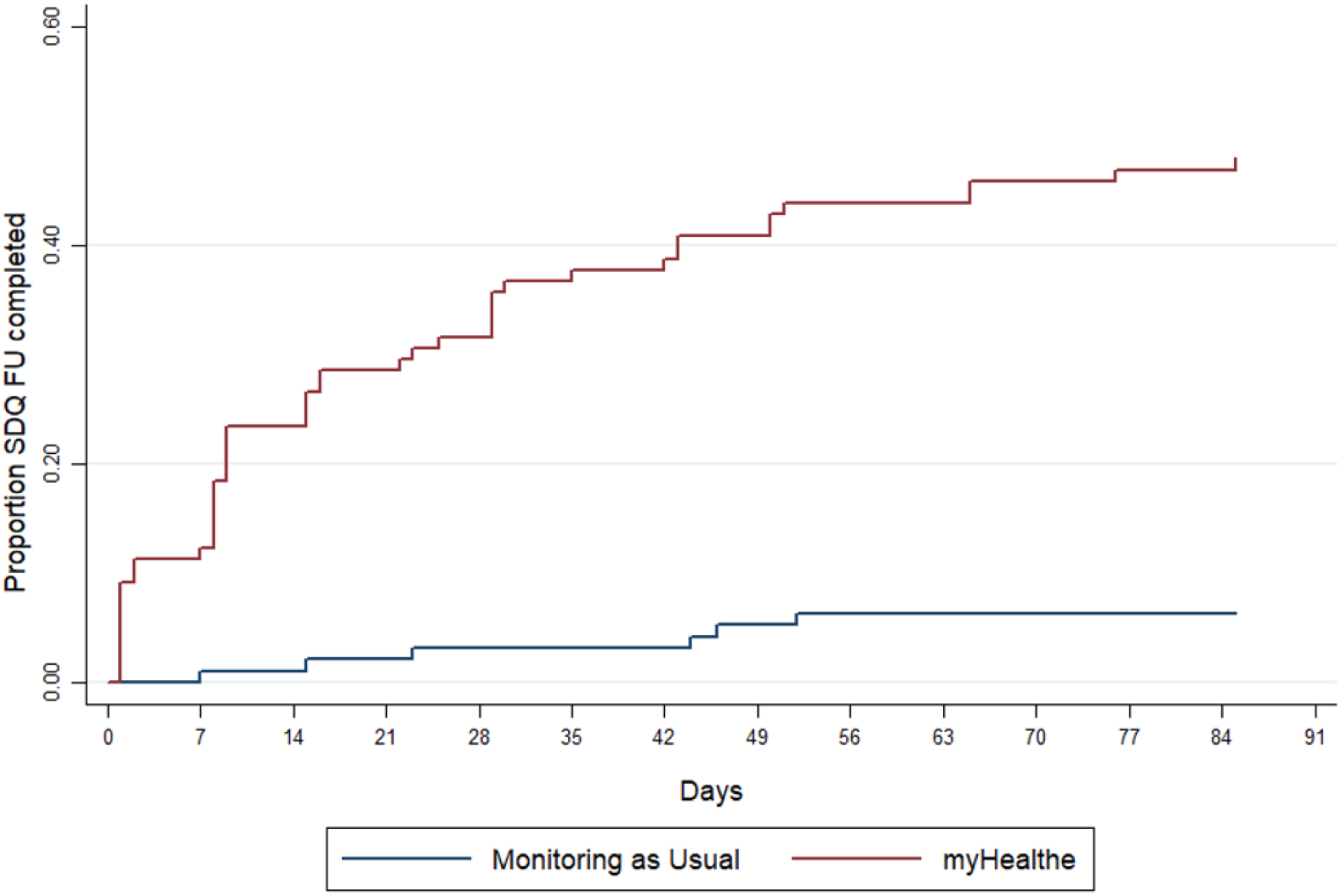
Kaplan-Meier curve illustrating the probability of SDQ-P within study period between caregivers assigned to complete electronic compared to paper SDQ-P.

### Caregiver perspective of MHE implementation

A total of eight SUS questionnaires and usability interviews were completed. The mean SUS score for users of the website was 78/100 indicating that the application was ‘acceptable’ to users. Figure 5. provides a summary of caregiver’s comments regarding MHE.

**Figure 5.**
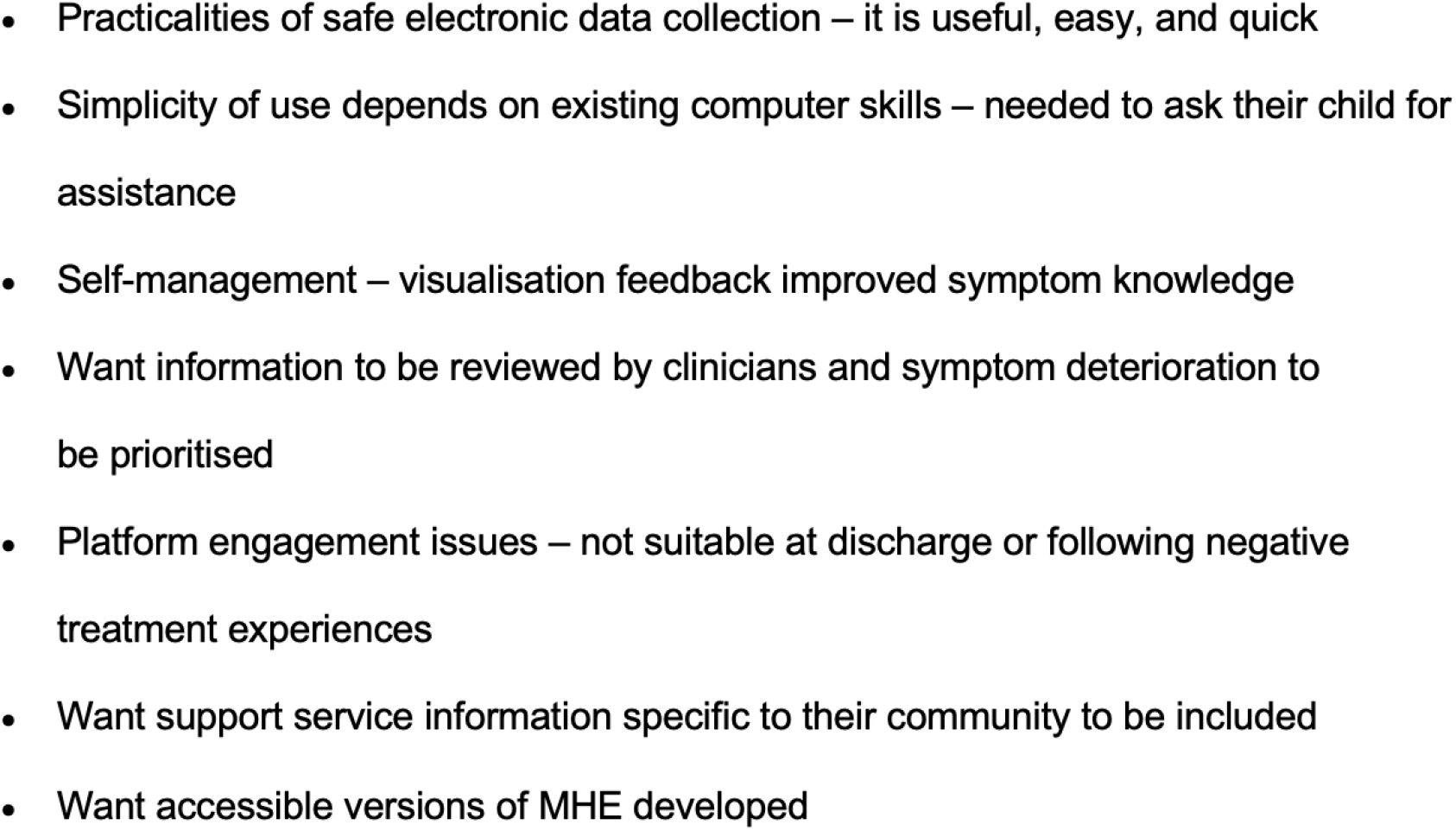
Summary of patient feedback following MHE use.

## Discussion

This trial showed that the collection of electronic PROMs using web-based technology is feasible and beneficial to CAMHS practice. Caregiver’s who received regular invitations to register for MHE had a SDQ-P completion rate of 69% compared to 12% paper-based SDQ-P completion. These finding represents a 61% absolute improvement, or 12 fold increase, of paired SDQ-P collection rates (Morris et al., 2020). By automating unassisted delivery of PROMs with completion prompts at specified time points, MHE addresses several fundamental challenges inherent to paper-based information gathering in busy clinical settings, such as processing burden, lack of supportive infrastructure and poor administration guideline knowledge (Waldron, Loades & Roadgers, 2018; Boswell et al. 2015; Duncan and Murray 2012; Wolpert, 2014).

In post-trial interviews caregivers rated MHE as ‘acceptable’, suggesting good levels of usability. Many caregivers favoured the ease and speed of using MHE to complete outcome measures compared to paper-based methods, while barriers included how readily information provided through the platform was used by clinicians to identify children with worsening symptoms and data privacy concerns. However, only a small number of caregivers were contacted to provide their views on the system; therefore, it is possible that other undetected usability issues influenced the results of this trial, for example: language, literacy level, disability, and cultural sensitivity difficulties (Lindsay, Bellaby, Smith et al., 2008; Morey, 2007; Kontos, Bennett, Viswanath, 2007; Bodie & Dutta, 2008).

Historically, low engagement with eHealth has been attributed to unequal internet access (Latulippe, Hamel & Giroux, 2017) but did not appear to account for non-engagement in the current trial. This finding is likely to reflect the substantial increase in mobile phones and other internet-enabled mobile technology availability (Pew Research Center, 2019), reduced cost of internet subscriptions and widening availability of free public Wi-Fi (Kontos et al., 2007; McAuley, 2014). However, despite physical internet access, end-users may not have the skills necessary to fully engage with digital technologies (Hargittai, 2002). This was the case for several caregivers who reported that their limited information technology capabilities and knowledge making it hard to navigate MHE without assistance from family members.

This disparity may deepen as digital platforms are increasingly integrated into routine clinical practice (Van Dijk, 2005) and should be iteratively considered during the design and implementation of emerging digital health platforms, paying particular attention to the role of co-design (Andersen, 2019).

## Strengths and Limitations

This trial was conducted in a naturalistic manner independent of clinical practice to ensure that clinician’s behaviour, for example, promoting MHE use did not inflate observed rates of engagement. Moreover, the research was conducted in a socio-demographically diverse geographical area, resulting in a broad range of caregivers testing the system. Finally, condition allocation was computerised meaning that all participants were instantly allocate to either receive MAU or MHE. Therefore, it was unlikely that allocation bias would have influenced the trial findings.

Limitations include the fact that families only had the opportunity to enrol to the trial if they had a baseline SDQ present in their child’s EHR, which relies on this being initiated by a clinician in the first instance. In the future, using MHE to capture baseline and follow up SDQ-P data may afford a more realistic assessment of ePROM feasibility. It is also possible that neurodevelopmental team service users perceived the SDQ-P as less useful than a disorder specific questionnaire, which may have resulted in lower rates of completion level. Lastly, owing to resource constraints phone interviews were conducted after the trial ended meaning that responses could be influenced by recall bias.

## Future research and MHE refinement

The next phase of this research is to extend this feasibility study across multiple-healthcare sites and other child mental health specialties and additional pertinent PROMs. Plans are already in place to extend MHE introduction to national and specialist teams and further SLaM CAMHS teams across Southwark, Lambeth, and Croydon. Recent funding secured from the National Institute for Health Research (NHIR; https://fundingawards.nihr.ac.uk/award/RP-PG-0618-20003) and the Medical Research Council (MRC) Mental Health Pathfinder award to King’s College London has enabled MHE to be converted into a scalable NHS software as a service (SaaS) product, with a roadmap to implement MHE across four other Trusts in England. Collecting data from a larger number of caregivers will enable us to explore the effects of various patient factors on ePROM engagement. Research investigating differential uptake in PROM collection suggests that several patient characteristics including ethnicity and social deprivation are associated with inequitable PROM use (Latulippe Hamel & Giroux, 2017, Morris et al., 2020). While this was not the case in the current small-scale trial, it is essential that further research is conducted to determine whether these systems sustain possible health inequalities with larger sample sizes.

In-depth interviews are needed to explore how ePROM platforms can be adapted to meet different service user needs and provide more general insights into caregivers’ reasons for deciding to complete or not complete electronic questionnaires, their views on the concept, design and delivery of MHE, the barriers and facilitators for MHE implementation and identify potential harms and study protocol refinement (e.g., platform design and frequency of questionnaire completion). System refinements are also required to enable alternative methods for acquiring and inputting caregiver contact information to circumvent the difficulties encountered with automatic data extraction in this study.

## Conclusion

Routine PROM collection is essential for delivering personalised health services that reflect clinical need from the perspective of young people and their families. This study supports the feasibility of a remote PROM monitoring platform within a real-world outpatient setting providing treatment to a demographically diverse population. Intimating that web-platforms may provide an acceptable and convenient method to maintain and scale up improved patient monitoring, service-user communication, and service evaluation. A future multisite trial of MHE is required to evaluate this e-system at scale.

## Supporting information

Supplemental Table 1, 2 and funding statement

## Data Availability

The data accessed by CRIS remain within an NHS firewall and governance is provided by a patient-led oversight committee. Access to data is restricted to honorary or substantive employees of the South London and Maudsley NHS Foundation Trust and governed by a local oversight committee who review and approve applications to extract and analyse data for research. Subject to these conditions, data access is encouraged and those interested should contact RS, CRIS academic lead.

## Acknowledgments

See online supplement 3.

## Appendix

### Appendix 1) Strengths and Difficulties Questionnaire

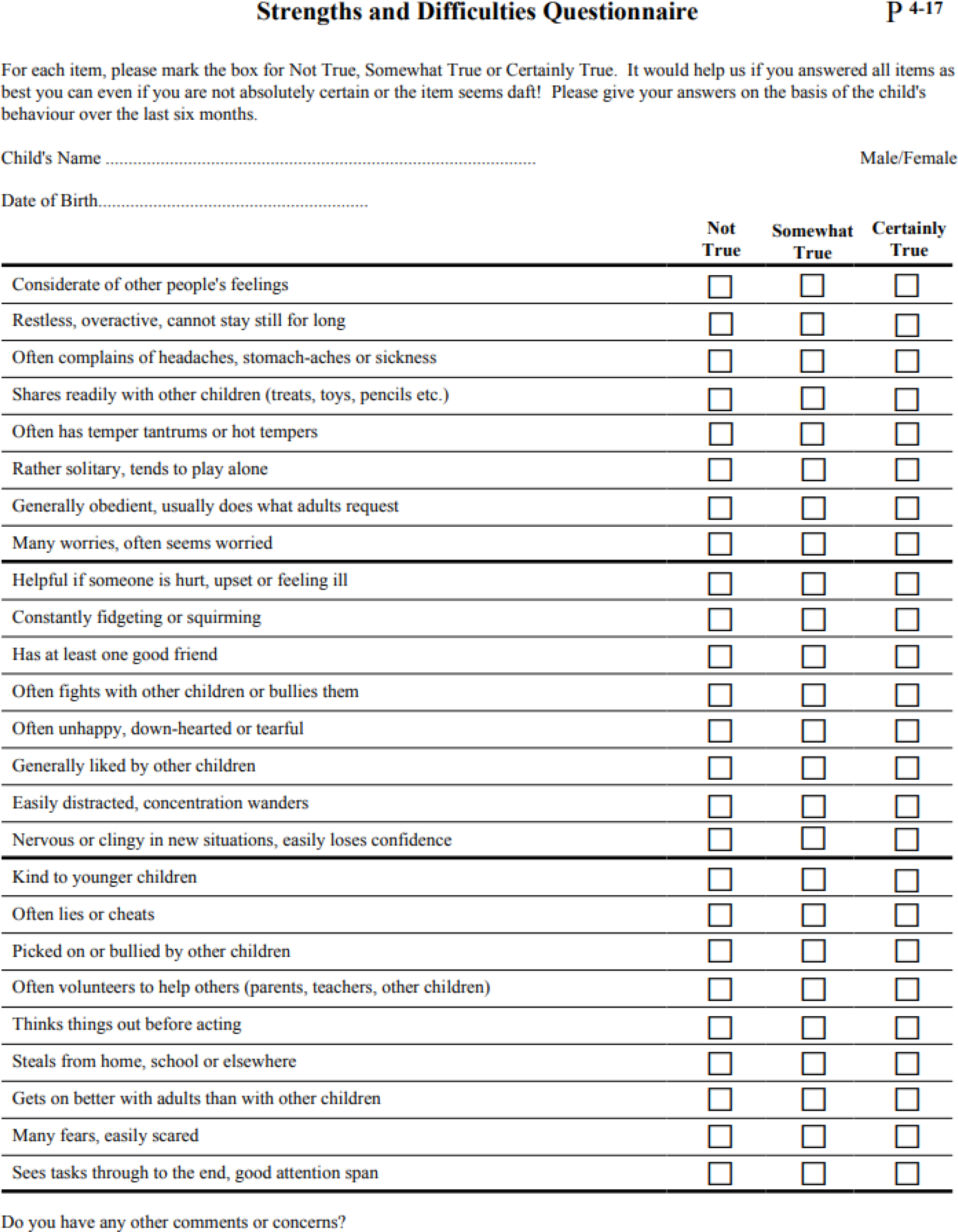

### Appendix 2a) MHE invitation text message; 2b) MHE reminder text message

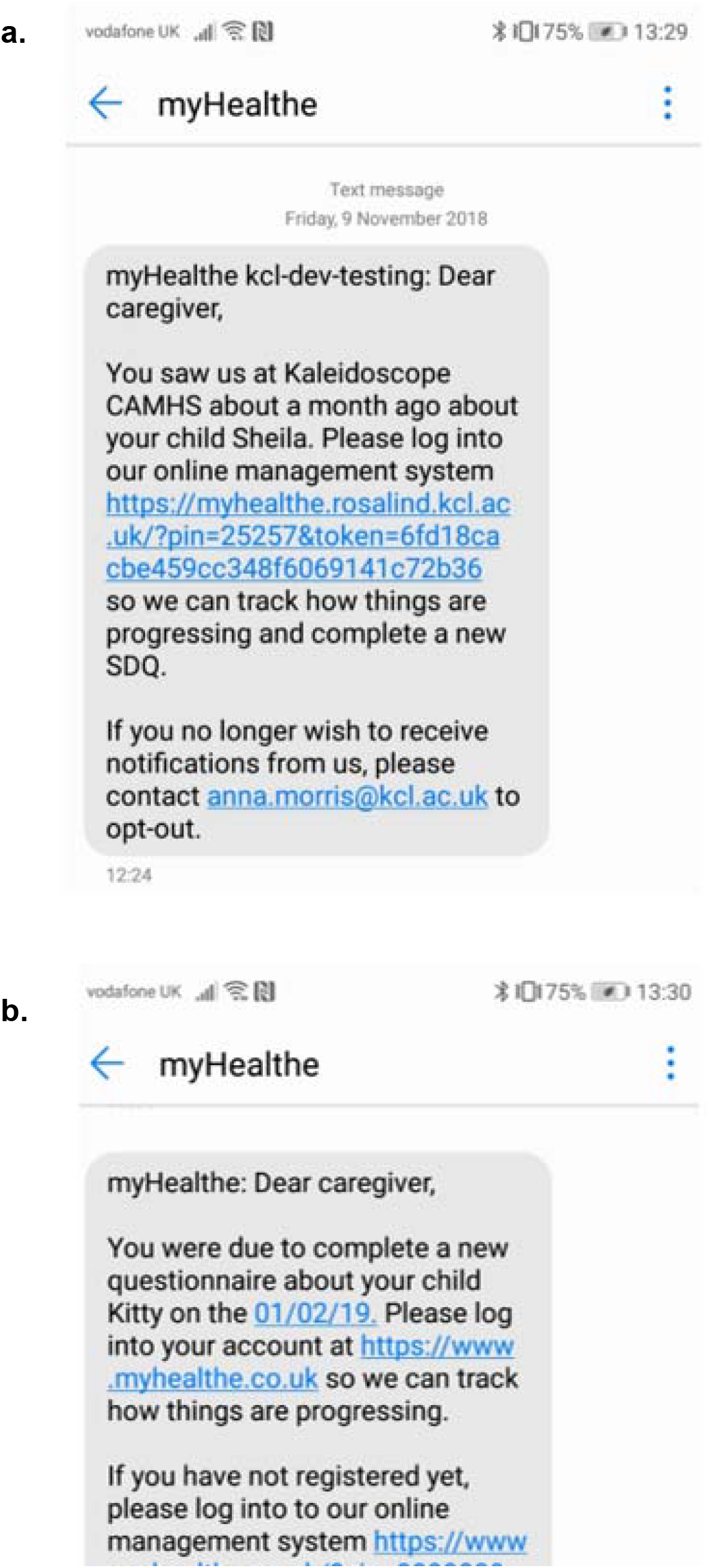

### Appendix 3a) MHE invitation email; 3b) MHE reminder email

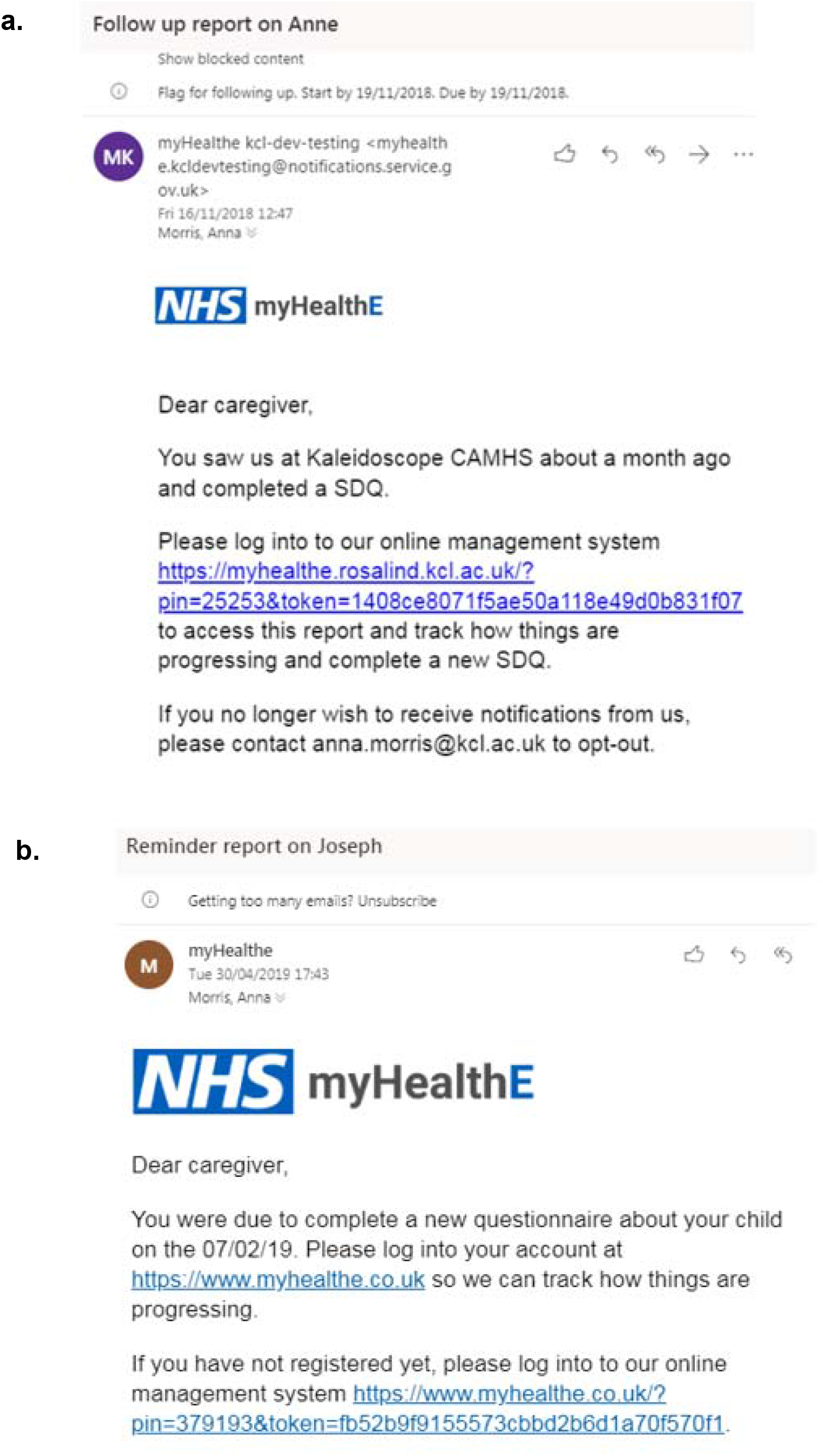

### Appendix 4) MHE login page - contact the corresponding author to request access to this image

### Appendix 5) MHE home page (when questionnaire is due to be completed - contact the corresponding author to request access to this image

### Appendix 5b) Electronic Strengths and Difficulties Questionnaire

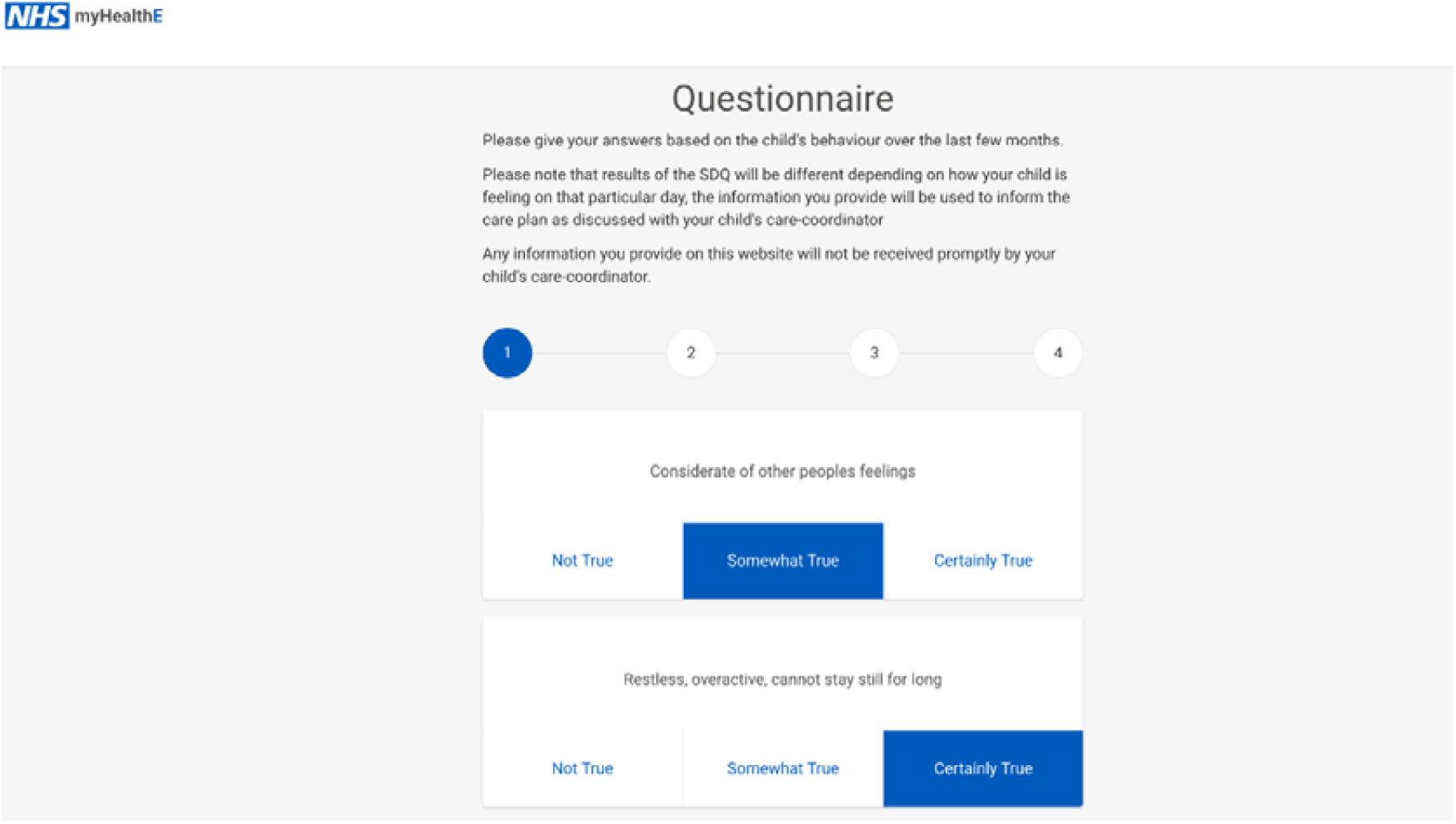

### Appendix 6a) Strengths and Difficulties Questionnaire results summary

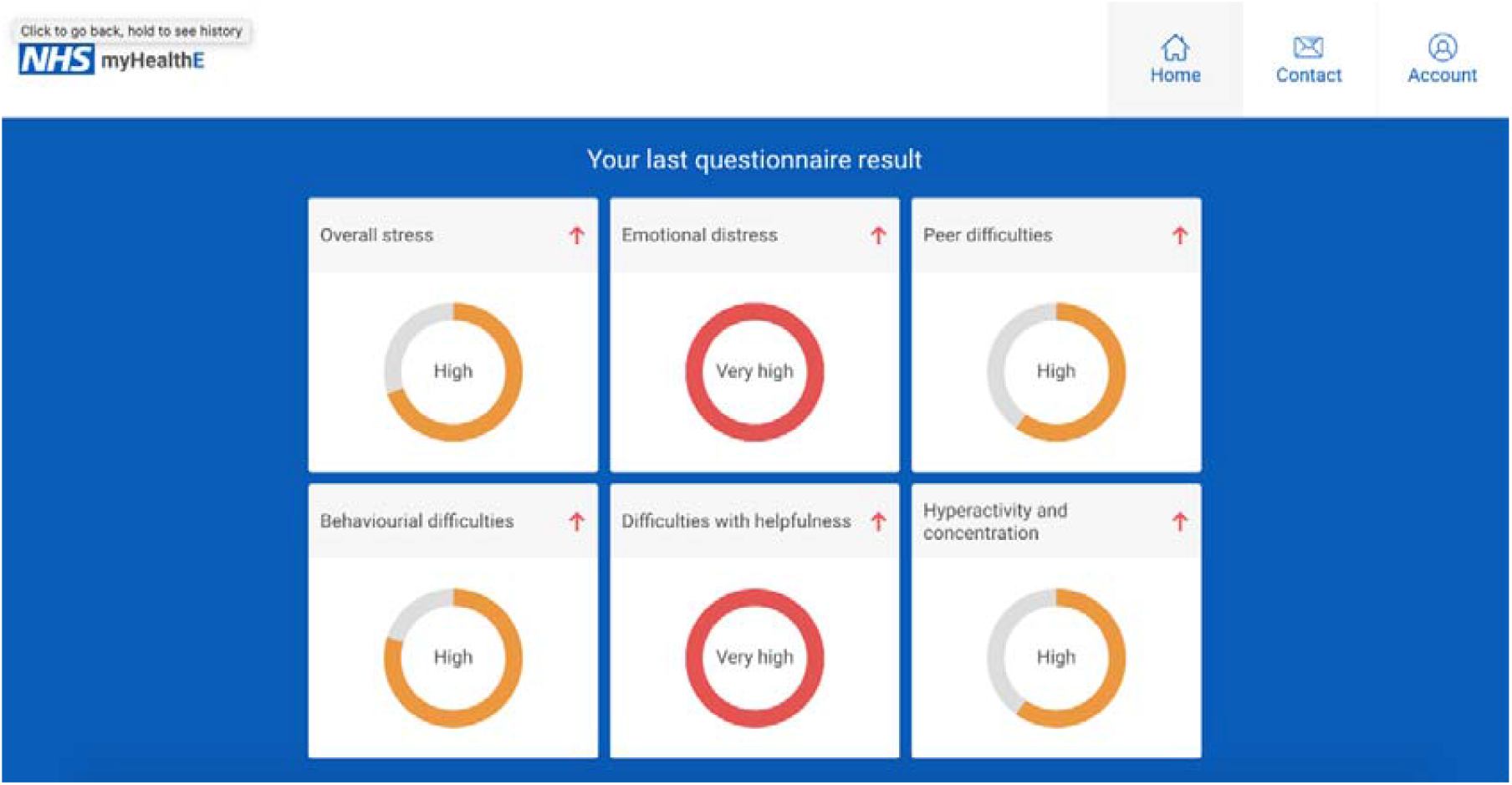

### Appendix 6b) Strengths and Difficulties Questionnaire previous results summary

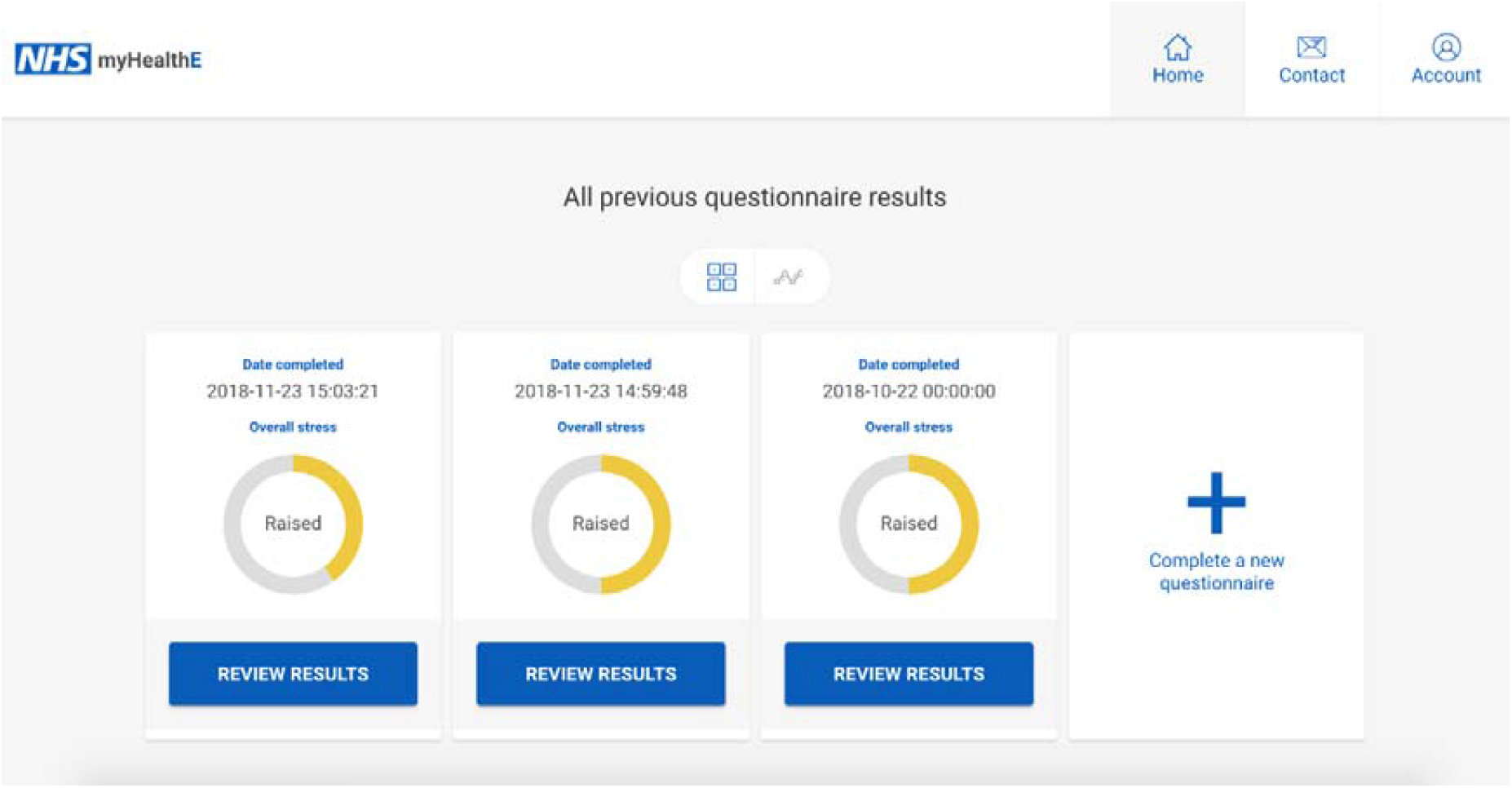

### Appendix 6c) Strengths and Difficulties Questionnaire results visualisation

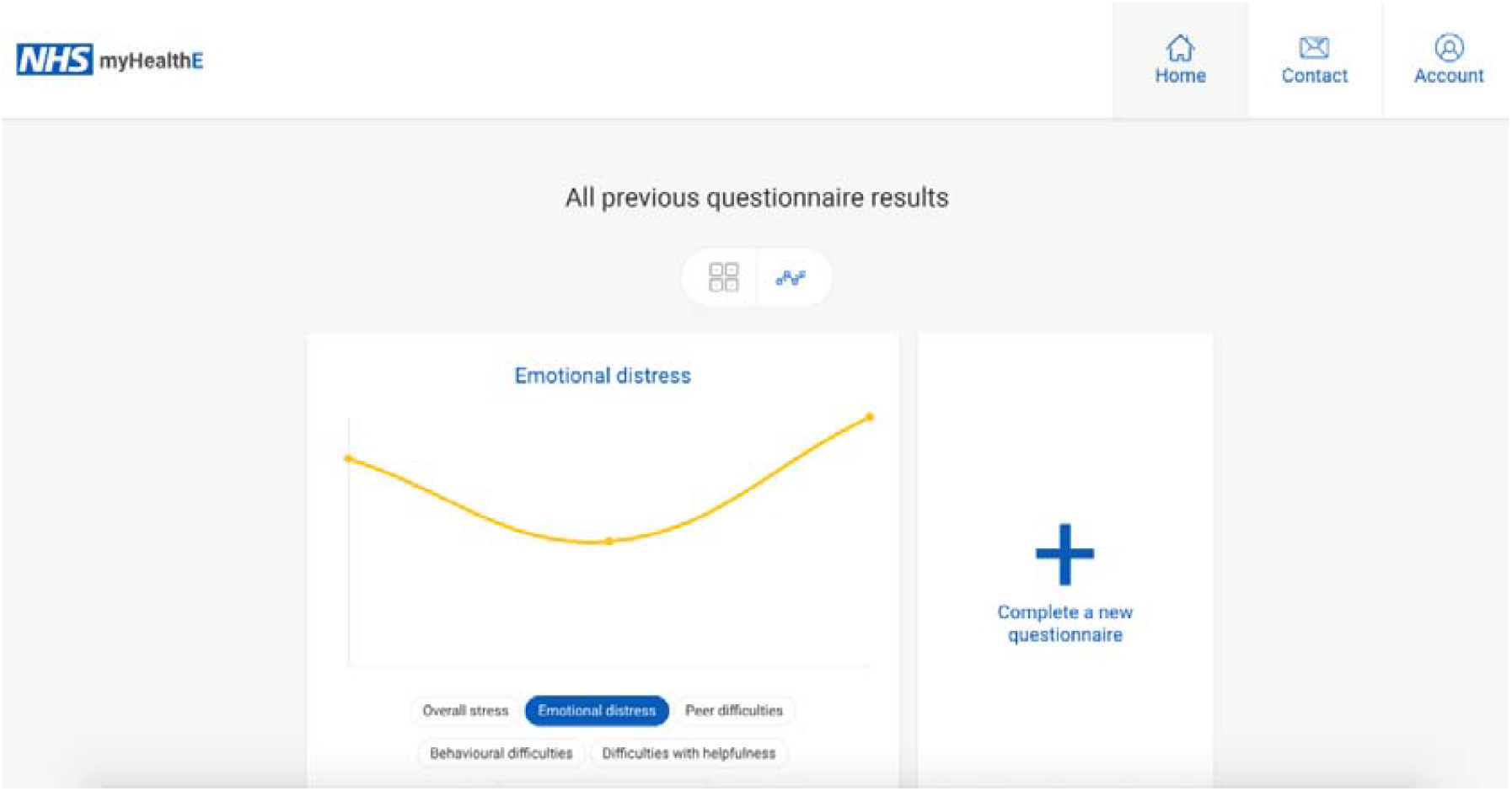

### Appendix 7a) MHE feasibility trial caregiver information sheet

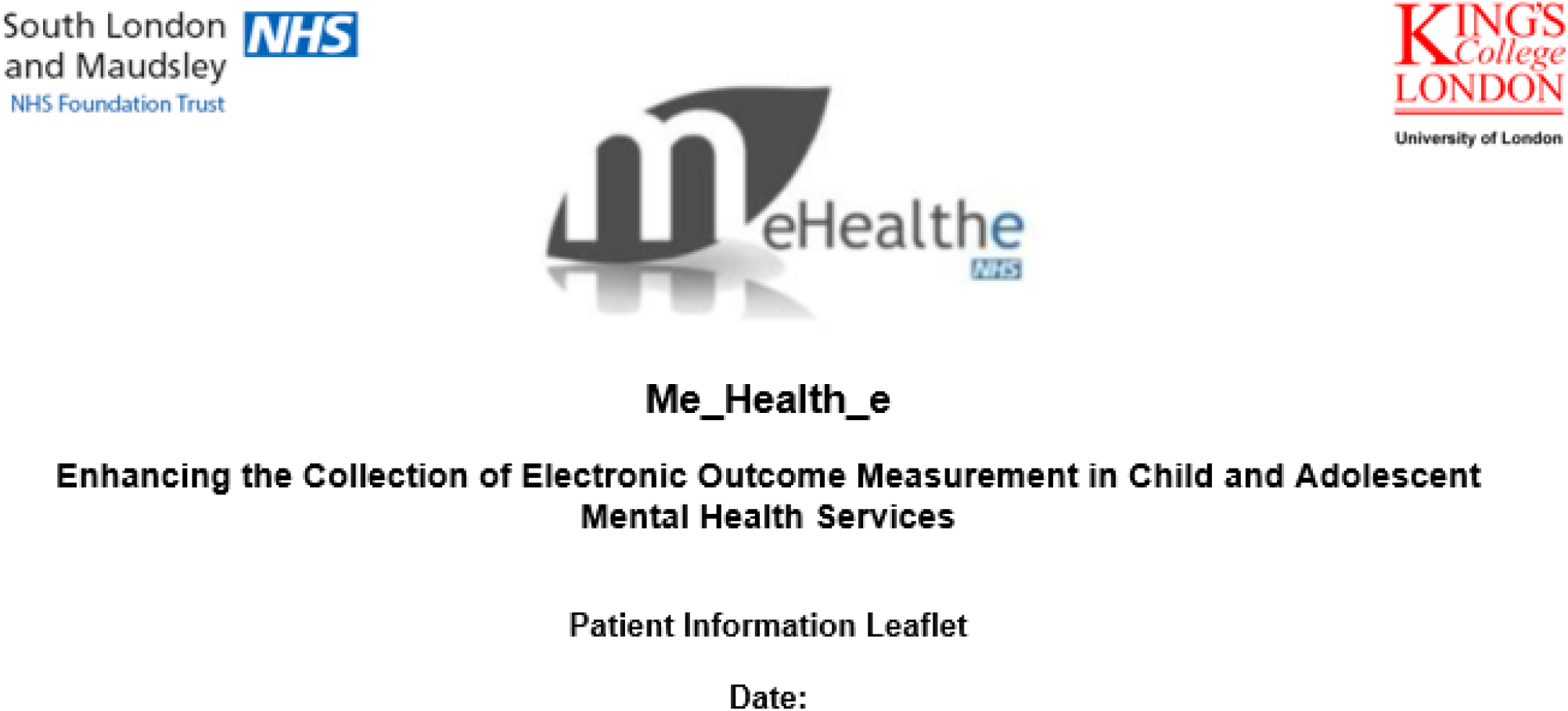

#### Overview

We are conducting an evaluation of an NHS online questionnaire system called Me_Health_e. This system aims to improve how child and adolescent mental health services (CAMHS) monitor changes in children and young people’s emotional and behavioural difficulties over time. The evaluation is being conducted by South London and Maudsley Hospital Foundation Trust (SLaM). Below we explain why this evaluation is being done and what your participation involves. If you have any questions, please do not hesitate to speak to your CAMHS worker or please get in contact with member of the evaluation team whose details are provided at the end of this information sheet.

#### Collecting Routine Outcome Measures

CAMH services are required to gather clinical information via questionnaires about your child from their first appointment until their discharge. These are called routine outcome measurements. Collecting this information allows our services to monitor your child’s progress, personalise care and ensure that you and your child’s opinions are accurately recorded when making decisions about care.

#### Why is this evaluation being conducted?

We are trying to improve the way we collect routine outcome measures in CAMHS. In particular we want to make it easier for you to provide information to clinicians. At the moment, we ask you to complete a 25 item paper based questionnaire called the Strengths and Difficulties Questionnaire (SDQ). We normally ask for this questionnaire to be completed in your first few appointments with CAMHS. After this, we ask for these questionnaires to be completed every 3-6 months or at the end of treatment. At present, these questionnaires are often not completed. Most CAMH services (in SLaM and across the UK) report that only 30% of the children they see, have 1 or more questionnaires collected at follow up.

In this evaluation we are testing the added value of an electronic system called the Me_Health_e to collect the SDQ. The Me_Health_e system works by automatically detecting when you are due to complete a SDQ and sends you an email or text reminder to complete an online version of the questionnaire.

#### How is this system being evaluated?

To understand whether Me_Health_e improves how we do things currently, we need to compare our standard way of collecting the SDQ with the Me_Health_e system. In order to do this, you will either continue with the standard method of collecting SDQ information or will be selected to complete the online survey. Whether you are allocated to conventional questionnaire completion or the online survey your child receives exactly the same mental health care.

#### Why have I been invited to take part?

You have been invited to take part because you have at least one child attending kaleidoscope either with the Lewisham CAMHS or Neurodevelopmental Team (NDT) and you have already have completed a paper based SDQ around your child’s first assessment with us.

#### What are the possible benefits of taking part?

We would encourage you to take part, as the information you provide does have an impact on how we track your child’s symptoms and allows us to measure the effectiveness of the CAMHS services as a whole.

If you choose to opt out and not complete the online SDQ it will not affect the level of care your child will receive. You will then be invited to complete SDQs in the standard way. If decide to take part and change your mind at a later date, you will still be able to withdraw yourself without reason and care for your child will remain unchanged.

#### What will my participation involve?

If you are asked to complete the SDQ online via the Me_Health_e system, we will need to have record of your most up to date email and/or mobile phone number. If these details are not already recorded in your child’s electronic health records, a member of our team will ask you to provide an email address and mobile phone number that you are happy to be contacted on. You can choose whether you would rather the Me_Health_e system contacts you on your email address or mobile phone number. Regardless of whether you are asked to complete a paper or online questionnaire, you will have already been asked or will be first asked to complete a paper SDQ. This will happen during your first session or shortly after. The number of times you will be asked to complete a new SDQ in addition to your baseline SDQ depends on which group your child has been assigned to. The maximum amount of SDQs you will be asked to complete is 6 over 6 months (1 per month).

##### Paper questionnaire (current practice)

If you are chosen to receive conventional paper based SDQ completion, after completing a first pen and paper SDQ, you will receive a reminder via post, phone or face-to-face to fill in a new paper SDQ. If your child’s treatment comes to an end before this six month period you may be asked to complete the SDQ sooner than this.

##### Online questionnaire

If you are chosen to receive the SDQ online you will be assigned to one of four groups. In all of these groups you will be asked to create a personal account in the Me_Health_e system. To do this, you will be emailed instructions and a link to the Me_Health_e website; this email will also contain a unique username and password which will allow you to make an account.

The next step will vary according to which group your child has been assigned too.

- ***Group 1*** After completing the first SDQ for your child, every month you will receive an email or text asking you to fill in a follow-up SDQ, this message will contain a link which will take you to an online version of the measure, you will receive a weekly reminder until the SDQ is filled. This will happen for a period of six months and you ’will be provided with a brief, automatically generated feedback report for each SDQ you complete which will be saved to your Me_Health_e account for you to access at any time.
- ***Group 2:*** After completing the first SDQ for your child, every month you ’will receive an email or text asking you to fill in a follow-up SDQ, this email will contain a link which will take you to an online version of the measure, you will receive a weekly reminder until the SDQ is filled. This will happen for a period of six months and you will not receive the brief feedback for any of the SDQs you complete.
- ***Group 3:*** After completing the first SDQ for your child, you will receive one reminder to fill in a followup at the end of 6 months. You will receive a feedback report for both SDQs you complete, which will be saved to your Me_Health_e account for you to access at any time.
- ***Group 4*** After completing the first SDQ for your child, you will receive one reminder to fill in a new SDQ at the end of 6 months and you will not receive feedback for any of the SDQs you complete.

If you decide at any point that you no longer want to receive emails from Me_Health_e there is an opt out option in the Me_Health_e navigation toolbar which you can tick to remove your details from this system.

#### Who will see the information collected from me?

As per normal clinical care, the clinical team working with your child will have access to the report generated from your responses on the SDQ. You can ask to see or discuss this at any time with CAMHS worker. Any analysis that is conducted on the information collected in this evaluation will be anonymised and carried out by approved SLaM staff, under SLaM Information Governance protocols.

#### What are the possible risks of taking pa rtf where is my data being stored?

Participating in this evaluation will not affect the level of care we provide for your child. All information provided via the online SDQ will be stored and protected on a secure server within the South London and Maudsley firewall. It will be treated as confidential information in the same way all your child’s health care records are managed.

#### What will happen to the findings of the research evaluation?

The findings from this evaluation will help us understand whether we should extend this system across other SLAM CAMH services. The findings will also be presented in medical journals and at meetings and conferences with other health care professionals. We will keep you informed of any updates during the evaluation and send you a summary of our findings once the evaluation is complete. All information presented are under strict data protection and SLaM governance protocols. No information will be presented that breaches patient confidentiality or identifies any individuals.

#### Who can I talk to if I have any questions?

Please feel free to discuss any questions you have about the evaluation or your involvement with your clinician during your appointment or contact our project lead Dr Johnny Downs ( or 020 3228 8553) or project support officer Anna Morris.

#### Who has reviewed this service evaluation?

This service evaluation has received approved as a Service Evaluation and Quality Improvement Project by the SLaM Clinical Audit Ethical approval office.

#### Who is leading and funding the research?

The research is being led by Dr Johnny Downs a Clinical Research Training Fellow and Honorary Child and Adolescent Psychiatrist at South London and Maudsley NHS Foundation Trust. The evaluation is being funded by Guy’s and St Thomas’ Charity Health Innovation Fund.

**Thank you for taking the time to read this information sheet and we thank you in advance for your help with this evaluation**.

### Appendix 7b) MHE feasibility trial caregiver leaflet - contact the corresponding author to request access to this image

## Notes

**Conflict of Interest:** None to declare.

### Competing Interest Statement

The authors have declared no competing interest.

### Clinical Trial

ISRCTN22581393

### Funding Statement

Z.I. M.P. R.S. M.H. A.P. R.J.B.D. E.S. and J.D. are affiliated with the National Institute of Health Research (NIHR) Biomedical Research Centre for Mental Health (BRC) Nucleus at the South London and Maudsley (SLaM) NHS Foundation Trust and Institute of Psychiatry Psychology and Neuroscience (IoPPN) Kings College London (KCL). Additionally the Clinical Record Interactive Search (CRIS) is supported by the NIHR BRC at the SLaM NHS Foundation Trust and KCL. A.C.M is supported by the Guys and St Thomas (GSST) Charity. Z.I. and R.J.B.D. are additionally supported by the NIHR University College London Hospitals BRC. R.J.B.D. is further supported by (1) Health Data Research (HDR) UK and (2) The BigData@Heart Consortium under grant agreement No. 116074. M.H. reports funding from the NIHR. A.S. is supported by the Intramural Research Program of the National Institute of Mental Health National Institutes of Health (NIH) (Grant No. ZIA-MH002957-01). R.S. is additionally part-funded by i) a Medical Research Council (MRC) Mental Health Data Pathfinder Award to Kings College London; ii) an NIHR Senior Investigator Award; iii) the NIHR Applied Research Collaboration South London (NIHR ARC South London) at Kings College Hospital NHS Foundation Trust. The views expressed are those of the authors and not necessarily those of the NIHR or the Department of Health and Social Care. M.H. declares funding from the Innovative Medicines Initiative for the RADAR-CNS consortium which includes contributions from Janssen MSD UCB Biogen and Lundbeck. A.P. is partially supported by the NIHR (NF-SI-0617-10120). E.S. is supported from the NIHR BRC at SLaM NHS Foundation Trust (IS‐BRC‐1215‐20018) the NIHR through a programme grant (RP‐PG‐1211‐20016) and Senior Investigator Award (NF‐SI‐0514‐10073 and NF‐SI‐0617‐10120) the European Union Innovative Medicines Initiative (EU‐IMI 115300) Autistica (7237) MRC (MR/R000832/1, MR/P019293/1) the Economic and Social Research Council (ESRC 003041/1) and GSST Charity (GSTT EF1150502) and the Maudsley Charity. J.D. is supported by NIHR Clinician Science Fellowship award (CS‐2018‐18‐ST2‐014) and has received support from a Medical Research Council (MRC) Clinical Research Training Fellowship (MR/L017105/1) and Psychiatry Research Trust Peggy Pollak Research Fellowship in Developmental Psychiatry.

### Author Declarations

Approval for the trial was given by the South London and Maudsley NHS Foundation Trust CAMHS Clinical Audit, Service Evaluation and Quality Improvement Committee (approval date: 07/04/2017). Extraction and analysis of deidentified outcome data were carried out using the CRIS platform and security model approved by Oxford Research Ethics Committee C (reference 18/SC/0372)

## References

Ashley, L., Jones, H., Thomas, J., Newsham, A., Downing, A., Morris, E., et al. (2013). Integrating patient reported outcomes with clinical cancer registry data: A feasibility study of the electronic Patient-Reported Outcomes from Cancer Survivors (ePOCS) system. Journal of Medical Internet Research, 15(10):e230. doi:10.2196/jmir.2764

Andersen, T. O. (2019) Large-scale and long-term co-design of digital health. Interactions 26(5): 74–77. doi: 10.1145/3356252

Bangor, A., Kortum, P. T., & Miller, J. T. (2008). An empirical evaluation of the system usability scale. International Journal of Human-Computer Interaction, 24(6), 574–594. doi: 10.1080/10447310802205776

Barthel, D., Fischer, K., Nolte, S., Otto, C., Meyrose, A.-K., Reisinger, S., et al. (2016). Implementation of the Kids-CAT in clinical settings: A newly developed computer-adaptive test to facilitate the assessment of patient-reported outcomes of children and adolescents in clinical practice in Germany. Quality of Life Research, 25(3), 585–94. doi: 10.1007/s11136-015-1219-9.

Batty, M. J., Moldavsky, M., Foroushani, P. S., Pass, S., Marriott, M., Sayal, K., & Hollis, C. (2013). Implementing routine outcome measures in child and adolescent mental health services: From present to future practice. Child and Adolescent Mental Health, 18(2), 82–87. doi: 10.1111/j.1475-3588.2012.00658.x

Black, M. M., & Ponirakis, A. (2000). Computer-administered interviews with children about maltreatment methodological, developmental, and ethical Issues. Journal of Interpersonal Violence, 15 (7), 682–695. doi: 10.1177/088626000015007002.

Black, N. (2013). Patient reported outcome measures could help transform healthcare. British Medical Journal, 346. doi: 10.1136/bmj.f167.

Bodie, G. D., & Dutta, M. J. (2008). Understanding health literacy for strategic health marketing: eHealth literacy, health disparities, and the digital divide. Health Marketing Quarterly, 25, 175–203. doi: 10.1080/07359680802126301.

Borzekowski, D. L. G., Leith, J., Medoff, D. R., Potts, W., Dixon, L. B., Balis, T., Hackman, A. L., & Himelhoch, S. (2009). Use of the Internet and other media for health information among clinic outpatients with serious mental illness. Psychiatric Services, 60(9), 1265–1268.

Boswell, J. F., Kraus, D. R., Miller, S. D., & Lambert, M. J. (2015). “Implementing routine outcome monitoring in clinical practice: Benefits, challenges, and solutions.” Psychotherapy Research, 25(1), 6–19. doi:10.1080/10503307.2013.817696

Breitenstein, S. M., & Gross, D. (2013). Web based delivery of a preventive parent training intervention: A feasibility study. Journal of Child and Adolescent Psychiatric Nursing, 26(2), 149–157. doi:10.1111/jcap.12031

Brooke, J. (1996). SUS: A “quick and dirty” usability scale. In P. W. Jordan, B. Thomas, B. A. Weerdmeester, & I. L. McClelland (Eds.), Usability evaluation in industry (pp. 189–194). London: Taylor & Francis.

Cella, D. F., Hahn, E.A, Jensen, S. E., Butt, Z., Nowinski, C. J., Rothrock, N., Lohr, K. N. (2015). Patient-reported outcomes in performance measurement. Research Triangle Park. Retrieved from https://www.rti.org/rti-press-publication/patient-reported-outcomes

Coons, S. J., Eremenco, S., Lundy, J. J., O’Donohoe, P., O’Gorman, H., Malizia, W. (2015). Capturing patient-reported outcome (PRO) data electronically: The past, present, and promise of ePRO measurement in clinical trials. Patient, 8:301–9. doi:10.1007/s40271-014-0090-z

Cruz, L. F. D. L., Simonoff, E., McGough, J. J., Halperin, J. M., Arnold, L. E., & Stringaris, A. (2015). “Treatment of children with attention-deficit/hyperactivity disorder (ADHD) and irritability: Results from the multimodal treatment study of children with ADHD (MTA).” Journal of the American Academy of Child & Adolescent Psychiatry, 54(1), 62–70.e3. doi:10.1016/j.jaac.2014.10.006.

Department of Health. (2004). National service framework for children, young people and maternity services: Core standards. Retrieved from: https://www.gov.uk/government/publications/national-service-framework-children-young-people-and-maternity-services

Dillon, D. G., Pirie, F., Rice, S., Pomilla, C., Sandhu, M. S., Motala, A. A., Young, E. H., & African Partnership for Chronic Disease Research (APCDR). (2014). Open-source electronic data capture system offered increased accuracy and cost-effectiveness compared with paper methods in Africa. Journal of clinical epidemiology, 67(12), 1358–1363. https://doi.org/10.1016/j.jclinepi.2014.06.012

Downs, J., Ford, T., Stewart, R., Epstein, S., Shetty, H., Little, R., et al. (2019). An approach to linking education, social care and electronic health records for children and young people in South London: A linkage study of child and adolescent mental health service data. BMJ Open, 9, e024355. doi:10.1136/bmjopen-2018-024355

Duncan, E. A., & Murray, J. (2012). The barriers and facilitators to routine outcome measurement by allied health professionals in practice: a systematic review. BMC Health Services Research, 12, 96. doi: https://doi.org/10.1186/1472-6963-12-96

Ebert, J. F., Huibers, L., Christensen, B., & Christensen, M. B. (2018). Paper- or web-based questionnaire invitations as a method for data collection: Cross-sectional comparative study of differences in response rate, completeness of data, and financial Cost. Journal of Medical Internet Research, 23, 20(1): e24. doi: 10.2196/jmir.8353.

Eremenco, S., Coons, S. J., Paty, J., Eremenco. (2014). PRO data collection in clinical trials using mixed modes: Report of the ISPOR PRO mixed modes good research practices task force. Value Health, 17, 501–16.doi:10.1016/j.jval.2014.06.005

Fernandes, A. C., Cloete, D., Broadbent, M. T., Hayes, R. D., Chang, C. K., Jackson, R. G., Roberts, A., Tsang, J., Soncul, M., Liebscher, J., Stewart, R., & Callard, F. (2013). Development and evaluation of a de-identification procedure for a case register sourced from mental health electronic records. BMC Medical Informatics and Decision Making, 13, 71. doi: 10.1186/1472-6947-13-71.

Goodman, R. (1997). The Strengths and Difficulties Questionnaire: A research note. Journal of Child Psychology and Psychiatry, 38, 581–586. doi:10.1111/j.1469-7610.1997.tb01545.x

Hall, C. L., Moldavsky, M., Baldwin, L., Marriott, M., Newell, K., Taylor, J., et al. (2013). The use of routine outcome Measures in two child and adolescent mental health services: A completed audit cycle. BMC Psychiatry, 13, 270. doi: 10.1186/1471-244X-13-270

Hall, C. L., Taylor, J., Moldavsky, M., Marriott, M., Pass, S., Newell, K., et al. (2014). A qualitative process evaluation of electronic session-by-session outcome measurement in child and adolescent mental health services. BMC Psychiatry, 14, 113. doi:10.1186/1471-244X-14-113

Hargittai, E. (2002). Second-level digital divide: Differences in people’s online skills. First Monday, 7(4). Retrieved from http://firstmonday.org/article/view/942/864

Jamison, R. N., Raymond, S. A., Levine, J. G., Slawsby, E. A., Nedeljkovic, S. S., & Katz, N. P. (2001). Electronic diaries for monitoring chronic pain: 1-year validation study, 91(3), 277–285. doi: 10.1016/S0304-3959(00)00450-4.

Johnston, C., & Gowers, S. (2005). Routine outcome measurement: A survey of UK child and adolescent mental health services. Child Adolescent Mental Health, 10, 133–139. doi:10.1111/j.1475-3588.2005.00357.x

Kontos, E. Z., Bennett, G. G., Viswanath, K. (2007). Barriers and facilitators to home computer and internet use among urban novice computer users of low socioeconomic position. Journal of Medical Internet Research, 9(4), e31. doi: 10.2196/jmir.9.4.e31

Latulippe, K., Hamel, C., & Giroux, D. (2017). Social health inequalities and eHealth: A literature review with qualitative synthesis of theoretical and empirical studies. Journal of Medical Internet Research, 27, 19(4), e136. doi:10.2196/jmir.6731.

Lindsay, S., Bellaby, P., Smith, S., & Baker, R. (2008). Enabling healthy choices: Is ICT the highway to health improvement? Health (London), 12(3), 313–331. doi:10.1177/1363459308090051

Lyon, A. R., Lewis, C. C., Boyd, M. R., Hendrix, E., & Liu, F. (2016). Capabilities and characteristics of digital measurement feedback systems: Results from a comprehensive review. Administration and Policy in Mental Health and Mental Health Services Research, 43, 441–466. doi:10.1007/s10488-016-0719-4.

McAuley, A. (2014). Digital health interventions: Widening access or widening inequalities? Public Health, 128(12), 1118–1120. doi: 10.1016/j.puhe.2014.10.008

McLennan, D., Barnes, H., Noble, M. Davies, J., Garratt, E., & Dibben, C. (2011). The English indices of deprivation. Retrieved from https://www.gov.uk/government/statistics/english-indices-of-deprivation-2015

Morey, O. T. (2007). Digital disparities: The persistent digital divide as related to health information access on the internet. Journal of Consumer Health on the Internet, 11(4), 23–41. doi: 10.1300/J381v11n04_03

Morris, A. C., Ibrahim, Z., Moghraby, O. S., Stringaris, A., Grant, I. M., Zalwwski, L., McClellan, S., Moriarty, G., Simonoff, E., Dobson, R. J. B., & Downs, J. (2021). medRxiv. doi:https://doi.org/10.1101/2021.06.09.21257998

Morris, A. C., Macdonald, A., Moghraby, O., Stringaris, A., Hayes, R. D., Simonoff, E., Ford, T., & Downs, J. M. (2020). Sociodemographic factors associated with routine outcome monitoring: a historical cohort study of 28,382 young people accessing child and adolescent mental health services. Child Adolescent Mental Health, 16. doi: 10.1111/camh.12396.

National Health Service. (2015). Future in mind: Promoting, protecting and improving our children and young people’s mental health and wellbeing. Retrieved from https://www.gov.uk/government/publications/improving-mental-health-servicesfor-young-people

Niazkhani, Z., Toni, E., Cheshmekaboodi, M., Georgiou, A., & Pirnejad, H. (2020). Barriers to patient, provider, and caregiver adoption and use of electronic personal health records in chronic care: a systematic review. BMC Medical Informatics and Decision Making, 20(1):153. doi:10.1186/s12911-020-01159-1.

Nordan, L., Blanchfield, L., Niazi, S., Sattar, J., Coakes, C. E., Uitti, R., et al. (2018). Implementing electronic patient-reported outcomes measurements: challenges and success factors. BMJ Quality and Safety, 27(10):852–6. doi:10.1136/bmjqs-2018-008426

Perera, G., Broadbent, M., Callard, F., Chin-Kuo, C., Downs, J., Dutta, R., et al. (2016). Cohort profile of the South London and Maudsley NHS Foundation Trust Biomedical Research Centre (SLaM BRC) Case Register: Current status and recent enhancement of an electronic mental health record-derived data resource. BMJ Open, 6:e008721. doi:10.1136/bmjopen-2015-008721

Pew Research Center. (2019) “Smartphone Ownership Is Growing Rapidly Around the World, but Not Always Equally”. Retrieved from: https://www.pewresearch.org/global/2019/02/05/smartphone-ownership-is-growing-rapidly-around-the-world-but-not-always-equally/

Schepers, S., Sint Nicolaas, S., Maurice-Stam, H., Van Dijk-Lokkart, E., Van den Bergh, E., De Boer, N., et al. (2017). First experience with electronic feedback of the Psychosocial Assessment Tool in pediatric cancer care. Supportive Care in Cancer, 25(10), 3113–3121. doi: 10.1007/s00520-017-3719-3.

Shaffer, D., Gould, M., Brasic, J., Ambrosini, P., Fisher, P., Bird, H., & Aluwahlia, S. (1983). A children’s global assessment scale. Archives of General Psychiatry, 40, 1228–1231.

StataCorp. (2015). Stata Statistical Software: Release 14. College Station, TX: StataCorp LP.

Steele Gray, C., Gill, A., Khan, A.I., Hans, P.K., Kuluski, K., & Cott, C. (2016a). The electronic patient reported outcome tool: Testing usability and feasibility of a mobile app and portal to support care for patients with complex chronic disease and disability in primary care settings. JMIR mHealth uHealth, 4(2):e58. doi: 10.2196/mhealth.5331.

Stevens, S., & Pritchard, A. (2020). Third phase of NHS response to COVID-19. Retrieved from: https://www.england.nhs.uk/coronavirus/wp-content/uploads/sites/52/2020/07/Phase-3-letter-July-31-2020.pdf

Stewart, R., Soremekun, M., Perera., G., Broadbent, M., Callard Denis, M., et al. (2009). The South London and Maudsley NHS Foundation Trust Biomedical Research Centre (SLaM BRC) case register: Development and descriptive data. BMC Psychiatry, 9, 51. doi:10.1186/1471-244X-9-51

Van Dijk, J. A. G. M. (2005). The deepening divide: Inequality in the information society. London, UK: Sage.

Waldron, S. M., Loades, M. E., & Rogers, L. (2018). Routine outcome monitoring in CAMHS: How can we enable implementation in practice? Child Adolescent Mental Health, 23(4), 328–333. doi: 10.1111/camh.12260.

Wolpert, M. (2014). Uses and abuses of patient reported outcome measures (PROMs): Potential Iatrogenic impact of PROMs implementation and how it can be mitigated. Administration and Policy in Mental Health and Mental Health Services Research, 41(2), 141–145. doi: 10.1007/s10488-013-0509-1

Zuidgeest, M., Hendriks, M., Koopman, L., Spreeuwenberg, P., Rademakers, J. (2011). A comparison of a postal survey and mixed-mode survey using a questionnaire on patients’ experiences with breast care. Journal of Medical Internet Research, 13(3):e68. doi: 10.2196/jmir.1241.

