## Supplemental Table 1, 2 and funding statement for "Assessing the Feasibility of a Web-based Outcome Measurement System in Child and Adolescent Mental Health Services: myHealthE (MHE) a Randomised Controlled Feasibility Pilot Study"

**Online supplement 1.** List of socio-demographic and clinical variables extracted from CRIS.

| **CRIS variable** | **Description** |
| --- | --- |
| Gender | Gender of the child recorded in CRIS at the trial start date. |
| Age | Age of the child recorded in CRIS at the trial start date. |
| Ethnicity | Child’s ethnicity was collapsed from source EHR codes into five broad categories (as defined by the UK Office for National Statistics): 1) White (White British, Irish and Other White Background), 2) Black (African, Caribbean and Other Black), 3) Asian (Indian, Pakistani, Bangladeshi, Chinese and Other Asian), 4) Mixed and Other (White and Black Caribbean, White and Black African, White and Asian and Other Mixed and any other ethnic group), and 5) Not stated (ethnicity not provided). |
| Neighbourhood deprivation | An index of neighbourhood deprivation for the main caregiver residence comprised in the current sample, categorised into quartiles of multiple deprivation (McLennan, Barnes, Noble et al., 2011). |
| ICD-10 primary or secondary diagnosis | Child’s most recent ICD-10 primary or secondary diagnosis at the trial start date. |
| Children’s Global Assessment Scale (CGAS) | Most recent CGAS (Shaffer, Gould, Brasic et al., 1983) outcome, a clinician reported assessment of patient functioning reported as a single score from 0 (extremely impaired) to 100 (doing very well). |
| Active care days | Number of active care days (inpatient and outpatient) recorded in the two years before the trial start date. |
| Face-to-face events | Number of attended face-to-face CAMHS events recorded in the two years preceding the trial start date. |
| SDQ sub-scale scores | SDQ sub-scale scores for the most recently recorded SDQ before the start of the trial. |

**Online supplement 2.** Description of caregiver opt-out preferences and technical difficulties encountered at MHE registration.

| **Platform engagement Issue** | **Reason** | **Outcome** |
| --- | --- | --- |
| Opt-out (n=2) | Patient 1: Felt it was unnecessarily complicated to set up an account on MHE and did not believe that using MHE would help their child receive quicker treatment.  Patient 2: Their child was no longer accessing CAMHS. | Patient 1: Successfully opted-out and received no further communication from MHE.  Patient 2: Successfully opted-out and received no further communication from MHE. |
| Technical difficulties (n=2) | Patient 1 & 2: MHE login page was not auto populated with the caregiver’s unique user ID, rendering users unable to set up their account. | Patient 1 & 2: Registration link was resent, and the issue desisted. |

**Online supplement 3.** Full list of funding, acknowledgements, and contributions.

Z.I., M.P., R.S., M.H., A.P., R.J.B.D., E.S., and J.D. are affiliated with the National Institute of Health Research (NIHR) Biomedical Research Centre for Mental Health (BRC) Nucleus at the South London and Maudsley (SLaM) NHS Foundation Trust and Institute of Psychiatry, Psychology and Neuroscience (IoPPN), King’s College London (KCL). Additionally, the Clinical Record Interactive Search (CRIS) is supported by the NIHR BRC at the SLaM NHS Foundation Trust and KCL. A.C.M is supported by the Guy’s and St Thomas’ (GSST) Charity. Z.I. and R.J.B.D. are additionally supported by the NIHR University College London Hospitals BRC. R.J.B.D. is further supported by (1) Health Data Research (HDR) UK and (2) The BigData@Heart Consortium under grant agreement No. 116074. M.H. reports funding from the NIHR. A.S. is supported by the Intramural Research Program of the National Institute of Mental Health National Institutes of Health (NIH) (Grant No. ZIA-MH002957-01). R.S. is additionally part-funded by i) a Medical Research Council (MRC) Mental Health Data Pathfinder Award to King’s College London; ii) an NIHR Senior Investigator Award; iii) the NIHR Applied Research Collaboration South London (NIHR ARC South London) at King’s College Hospital NHS Foundation Trust. The views expressed are those of the authors and not necessarily those of the NIHR or the Department of Health and Social Care. M.H. declares funding from the Innovative Medicines Initiative for the RADAR-CNS consortium which includes contributions from Janssen, MSD, UCB, Biogen and Lundbeck. A.P. is partially supported by the NIHR (NF-SI-0617-10120). E.S. is supported from the NIHR BRC at SLaM NHS Foundation Trust (IS‐BRC‐1215‐20018), the NIHR through a programme grant (RP‐PG‐1211‐20016) and Senior Investigator Award (NF‐SI‐0514‐10073 and NF‐SI‐0617‐10120), the European Union Innovative Medicines Initiative (EU‐IMI 115300), Autistica (7237) MRC (MR/R000832/1, MR/P019293/1), the Economic and Social Research Council (ESRC 003041/1) and GSST Charity (GSTT EF1150502) and the Maudsley Charity. J.D. is supported by NIHR Clinician Science Fellowship award (CS‐2018‐18‐ST2‐014) and has received support from a Medical Research Council (MRC) Clinical Research Training Fellowship (MR/L017105/1) and Psychiatry Research Trust Peggy Pollak Research Fellowship in Developmental Psychiatry. The authors give thanks to the families and Kaleidoscope staff who participated in this trial. J.D conceived the trial aims, supervised data analysis and writing. A.C.M led on data analysis and manuscript writing. M.P. assisted with study design and data acquisition. All authors reviewed and provided critical revisions to the manuscript and approved the final version of the manuscript. The authors have no conflicts of interest to declare.
